# Mapping genetic convergence across brain structure, mental health, and cardiometabolic disease

**DOI:** 10.1101/2025.05.22.25328130

**Authors:** Jakub Kopal, Alexey A. Shadrin, Dennis van der Meer, Olav B. Smeland, Sara E. Stinson, Linn Rødevand, Nadine Parker, Kevin S. O’Connell, Oleksandr Frei, Srdjan Djurovic, Anders M. Dale, Ole A. Andreassen

## Abstract

Individuals with psychiatric disorders frequently experience comorbid cardiometabolic conditions, complicating treatment and worsening health outcomes. Both psychiatric and cardiometabolic disorders have been individually associated with alterations in brain structure. Yet, it remains unclear whether these associations reflect a shared genetic basis that also contributes to their frequent co-occurrence. Here we analyzed genome-wide association summary statistics for psychiatric disorders, cardiometabolic disease, and brain morphology using complementary genetic approaches to disentangle genetic factors underlying brain alterations and comorbidity. Our results highlighted differences in patterns of genetic overlap across disorders. Schizophrenia showed substantial polygenic overlap with cortical thickness and type 2 diabetes, despite low genetic correlation. In contrast, ADHD was more genetically correlated with cardiometabolic disease but showed limited overlap with cortical morphology. Mediation analysis suggested that cortical surface area may partly mediate the genetic link between ADHD and type 2 diabetes. Pathway enrichment highlighted metabolic stress in ADHD and neurodevelopmental and immune processes in schizophrenia. These findings suggest that psychiatric–cardiometabolic comorbidity arises through both shared and disorder-specific genetic pathways, clarifying the genetic architecture of multimorbidity and informing trait-targeted prevention strategies in psychiatry.

## Introduction

A whole-body perspective on mental health redefines the study of psychiatric disorders by emphasizing their integration with broader physiological systems^1^. Traditionally, mental health research has focused on the brain as the central organ of interest^2^. However, emerging evidence highlights how physical health, including cardiovascular function, immune responses, and metabolic pathways, contribute to the onset, progression, and manifestation of psychiatric disorders^3–6^. Given that most human diseases result from a complex interplay of multiple genetic variants and environmental factors^7^, exploring the genetic links between mental health, physical health, and brain organization offers a window into shared biological mechanisms. This multi-system perspective can pave the way for more effective treatment and prevention strategies.

Severe psychiatric disorders are known to be highly heritable, with twin studies estimating genetic contributions to vary between 40% and 80%^8–11^. Genome-wide association studies (GWAS) have advanced our understanding of the genetic complexity underlying mental disorders by identifying thousands of common genetic variants, each with an individually small effect^12^. Furthermore, novel analytical methods together with expanding genomic resources have revealed shared genetic architectures between distinct traits and disorders^13–15^. Specifically, studies indicate that multiple disorders may be linked by common genetic and biological pathways^16,17^. These pleiotropic mechanisms may contribute to the co-occurrence of conditions and call for a thorough examination of disease-specific vs. generalized patterns to better understand multimorbidity as well as the unique characteristics of each disorder.

Multimorbidity is an escalating global challenge^18^. Individuals with severe mental illness are more than twice as likely to experience physical multimorbidity^19–21^. These individuals often suffer from a range of physical health issues, including diabetes mellitus, hypertension, obesity or coronary heart disease, which further complicate diagnosis, treatment, and management^22^. Notably, all types of mental disorders are associated with significantly increased mortality rates^23^. Patients with schizophrenia (SCZ) have a life expectancy 15–20 years shorter than that in the general population where the higher mortality is predominantly due to cardiovascular and respiratory disease^24,25^. Similarly, Attention-Deficit/Hyperactivity Disorder (ADHD) has been associated with elevated risks of nervous system disorders (particularly sleep disorders), as well as cardiovascular, respiratory, musculoskeletal, and metabolic diseases^26,27^. While cardiometabolic diseases emerge as a common thread among several psychiatric disorders^28^, the potential underlying mechanisms for this comorbidity are mainly unknown. Several studies have implicated shared genetic factors between mental disorders and cardiometabolic traits^29–31^, yet the extent to which multimorbidity arises from shared genetic risk factors versus environmental influences remains a topic of ongoing investigations^32^. Recent results suggest that some links between cardiometabolic diseases and mental health disorders are primary, driven by shared biological mechanisms, while others are secondary, resulting from lifestyle factors, medication effects, or poor healthcare. The neurobiological mechanisms underlying the body-brain interplay thus remain unclear.

Brain alterations often mediate the effects of genetic variation on behavioral differentiation. Psychiatric disorders are accompanied by alterations in heritable regional brain metrics, including cortical thickness (CT) and surface area (SA)^33–37^. Recent analyses demonstrated extensive pleiotropy between psychiatric disorders and MRI-based measurements of CT as well as SA^16,38–40^. At the same time, studies have shown that brain morphometry is linked to cardiometabolic disorders^41,42^, with evidence suggesting a genetic basis for this association^43^. Despite numerous studies exploring brain MRI as a link between cardiometabolic disease and mental disorders, it remains unclear whether this relationship is disease-specific or reflects a generalized pattern shared across disorders. Clarifying this distinction could have significant clinical implications, particularly in determining whether the same therapeutic interventions can be applied across conditions. These insights are especially relevant for prevention and treatment efforts, as the gap in physical multimorbidity between those with and without severe mental illness is greatest in younger individuals, highlighting the need for early intervention^19^.

The growing availability of large-scale clinical, genetic, and neuroimaging data, combined with advanced analytical methods, offers new opportunities to dissect the shared genetic architecture of mental and physical health. We hypothesize that by simultaneously analyzing genetic overlap among psychiatric disorders, cardiometabolic disease, and cortical morphology, we can uncover whether the body-brain interplay underlying multimorbidity follows a disease-specific pattern or reflects a broader, cross-disorder mechanism shared across multiple conditions. For this purpose, we investigated genome-wide data from the five most prevalent psychiatric disorders—ADHD, autism spectrum disorder (ASD), major depressive disorder (MDD), bipolar disorder (BIP), and SCZ—together with average CT and total SA (following ^40^), as well as coronary artery disease (CAD) and type 2 diabetes (T2D). We focused on CAD and T2D as representative cardiometabolic diseases due to their high prevalence, consistent links to adverse brain outcomes, and complementary coverage of vascular and metabolic dysfunction^44,45^. We employed a comprehensive suite of genetic tools ranging from genetic correlation and polygenic overlap to causal inference, mediation analysis, and pathway enrichment. Notably, the novel trivariate MiXeR tool allowed us to deconstruct the patterns of polygenic overlap among three complex phenotypes that could not be deduced from bivariate methods^46^. Our results reveal both common and distinct genetic patterns across psychiatric disorders, emphasizing the heterogeneous genetic architecture that shapes their links to cardiometabolic health and brain structure.

## Results

### Genetic architectures of psychiatric and cardiometabolic diseases

We systematically analyzed the genetic overlap among psychiatric disorders, cardiometabolic diseases, and cortical morphology. To this end, we utilized respective GWAS summary statistics to quantify shared genetic architecture across these traits (Table 1). We begin by revisiting previous findings from our and other groups^16,40^, where LD score regression (LDSC) analysis revealed notable genetic distinctions among psychiatric disorders, laying the foundation for the current study. Specifically, ADHD and SCZ displayed the lowest genetic correlation among the investigated psychiatric disorders (r_G_ = 0.20; Fig. 1A). Furthermore, we observed striking differences in the genetic correlation of major mental disorders with cardiometabolic diseases represented by CAD and T2D (Fig. 1B). While ADHD and MDD displayed positive genetic correlations with CAD (ADHD: r_G_ = 0.30; MDD: r_G_ = 0.25), ASD, BIP, and SCZ showed correlations close to zero (ASD: r_G_ = −0.01; BIP: r_G_ = 0.03; SCZ: r_G_ = −0.03). A similar pattern of associations was observed for T2D (Fig. 1B). In addition, psychiatric disorders differed in their associations with cortical morphology represented by CT and SA. While all of them showed minimal genetic correlation with CT, ADHD and MDD exhibited negative correlations with SA (ADHD: r_G_ = −0.19, MDD: r_G_ = −0.10). The relatively low genetic similarity between SCZ and ADHD, coupled with their divergent associations with CAD and T2D, highlights differences in their genetic architecture and suggests that these disorders may be linked to cardiometabolic disease through distinct biological mechanisms (Fig. 1C). All phenotypes included in the analysis passed recommended quality control criteria, including adequate polygenicity and heritability to support advanced statistical modeling (Fig. 1D). Collectively, these findings indicate that SCZ and ADHD represent genetically distinct diagnoses that are well suited for further investigation into their shared and unique genetic links with cardiometabolic disease and cortical morphology.

**Figure 1:**
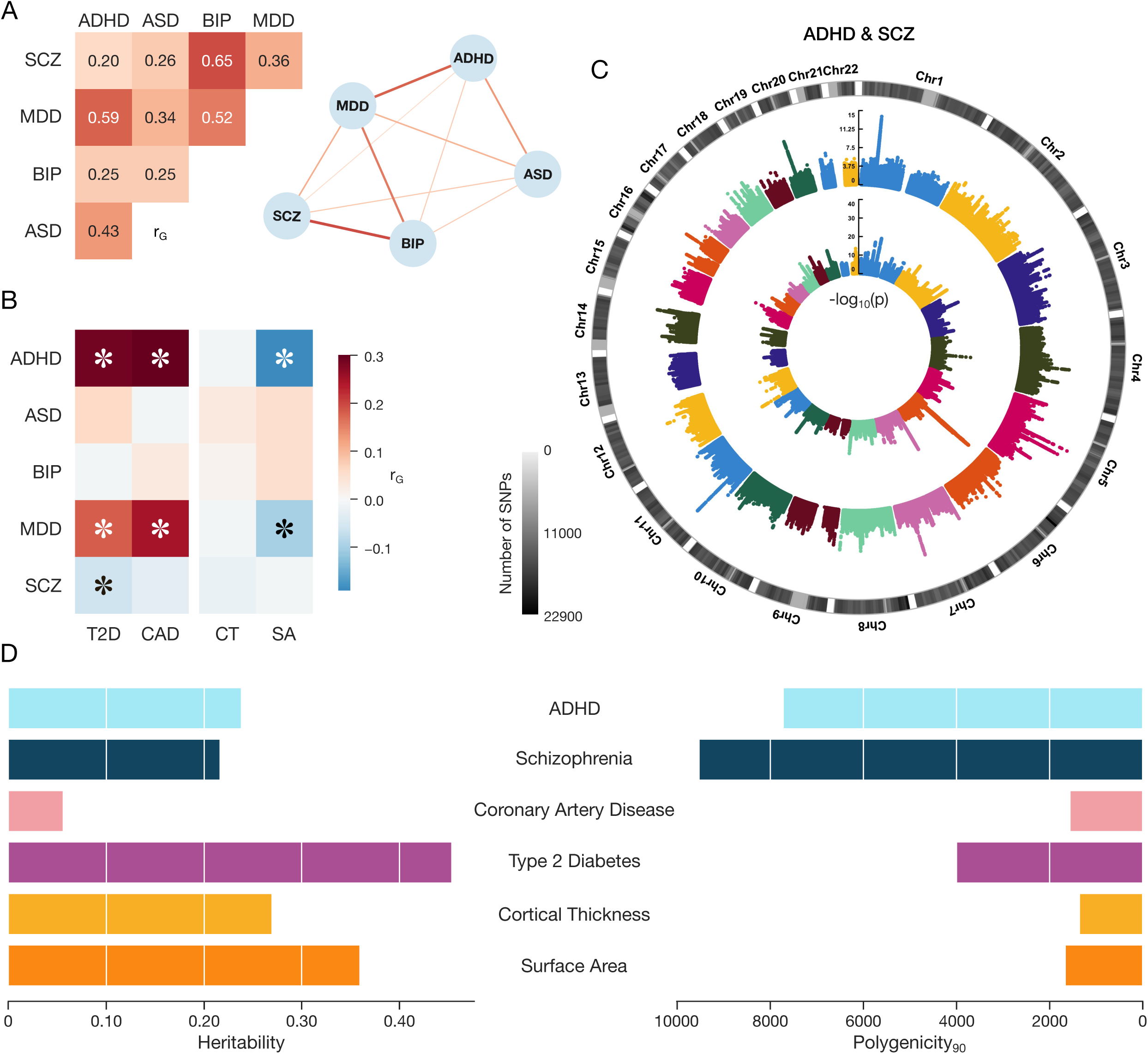
The distinct and shared genetic architecture of ADHD and SCZ. **A.** ADHD and SCZ show differences in genetic architecture. We calculated genetic correlations using LDSC between all pairs of the five most common psychiatric disorders. The resulting heatmap is then re-represented as a network diagram where each node is a psychiatric disorder and each edge corresponds to genetic correlation. **B.** Diverging relationship with cardiometabolic disease. The heatmap displays genetic correlations between the five psychiatric disorders and two representatives of cardiometabolic disease and cortical morphology, i.e., coronary artery disease (CAD) and type 2 diabetes (T2D) as well cortical thickness (CT) and surface area (SA). Statistically significant correlations after FDR correction are marked with asterisks. **C.** Genetic architecture of ADHD and SCZ. The outer ring shows the −log_10_(p-values) of SNP associations with ADHD, while the inner ring displays the −log_10_(p-values) for ADHD. The color gradient on the left represents SNP density across the genome. **D.** Heritability and polygenicity of analyzed traits. The bar plots display the estimated heritability estimated by LDSC and polygenicity estimated by univariate MiXeR of psychiatric disorders (SCZ, ADHD), cardiometabolic diseases (CAD, T2D), and cortical morphology (CT, SA). Heritability estimates for the disorders/diseases are presented on the liability scale, using population prevalence values consistent with those reported in ^16^. Polygenicity indicates the number (in thousands) of causal variants with the strongest effect sizes required to explain 90% of the SNP-based heritability. Collectively, the differences observed in genetic correlation analyses of SCZ and ADHD motivate further investigation into their genetic overlaps with cardiometabolic disease and cortical morphology.

**Table 1:**
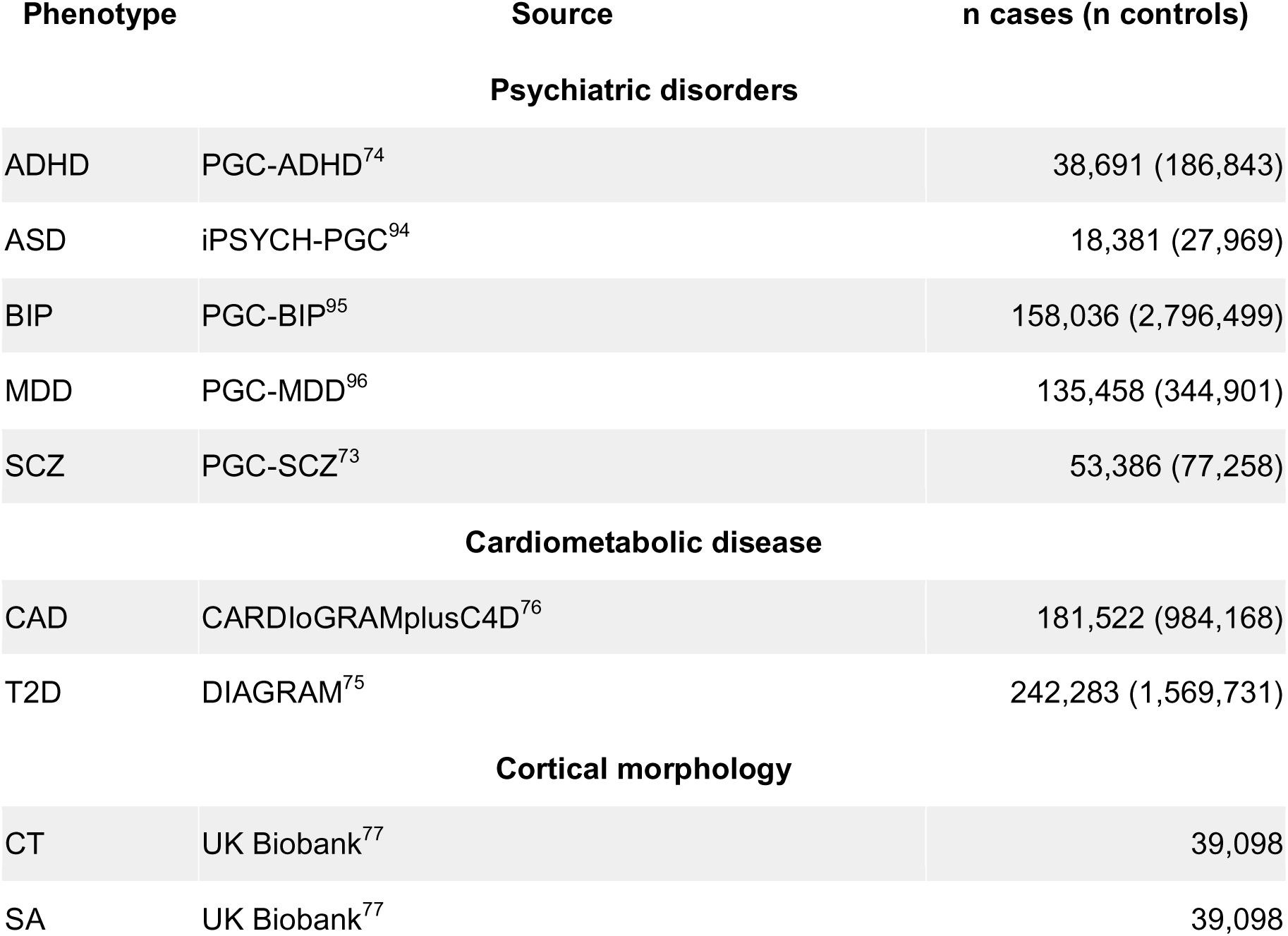
GWAS summary statistics. Overview of GWAS summary statistics used in the study. All analyses were restricted to individuals of European ancestry. Abbreviations: ADHD = attention-deficit/hyperactivity disorder, ASD = autism spectrum disorder, BIP = bipolar disorder, CAD = coronary artery disease, CT = cortical thickness, MDD = major depressive disorder, PGC = Psychiatric Genomics Consortium, SA = surface area, SCZ = schizophrenia, T2D = type 2 diabetes.

| Phenotype | Source | n cases (n controls) |
| --- | --- | --- |
| Psychiatric disorders |  |  |
| ADHD | PGC-ADHD<sup>74</sup> | 38,691 (186,843) |
| ASD | iPSYCH-PGC<sup>94</sup> | 18,381 (27,969) |
| BIP | PGC-BIP<sup>95</sup> | 158,036 (2,796,499) |
| MDD | PGC-MDD<sup>96</sup> | 135,458 (344,901) |
| SCZ | PGC-SCZ<sup>73</sup> | 53,386 (77,258) |
| Cardiometabolic disease |  |  |
| CAD | CARDIoGRAMplusC4D<sup>76</sup> | 181,522 (984,168) |
| T2D | DIAGRAM<sup>75</sup> | 242,283 (1,569,731) |
| Cortical morphology |  |  |
| CT | UK Biobank<sup>77</sup> | 39,098 |
| SA | UK Biobank<sup>77</sup> | 39,098 |

### Trivariate overlap between cardiometabolic disease, brain, and mental health

We leveraged trivariate MiXeR to directly estimate the genetic overlap between psychiatric disorders, cardiometabolic diseases and cortical morphology. Notably, the trivariate overlap was less pronounced for ADHD compared to SCZ. Given that ADHD exhibited stronger genetic overlap and genetic correlation with SA, we focused on presenting the overlaps between ADHD, SA, and cardiometabolic diseases (Fig. 2A). Results for CT are provided in Sup. Fig. 1. We found a similar degree of genetic overlap between ADHD and CAD (4%) and between ADHD and SA (4%), while fewer than 1% of trait-influencing variants were shared across all three traits. In contrast, the overlap between ADHD and T2D was higher (11%), compared to SA (4%), with an additional 1% of trait-influencing variants shared jointly among ADHD, SA, and T2D.

**Figure 2:**
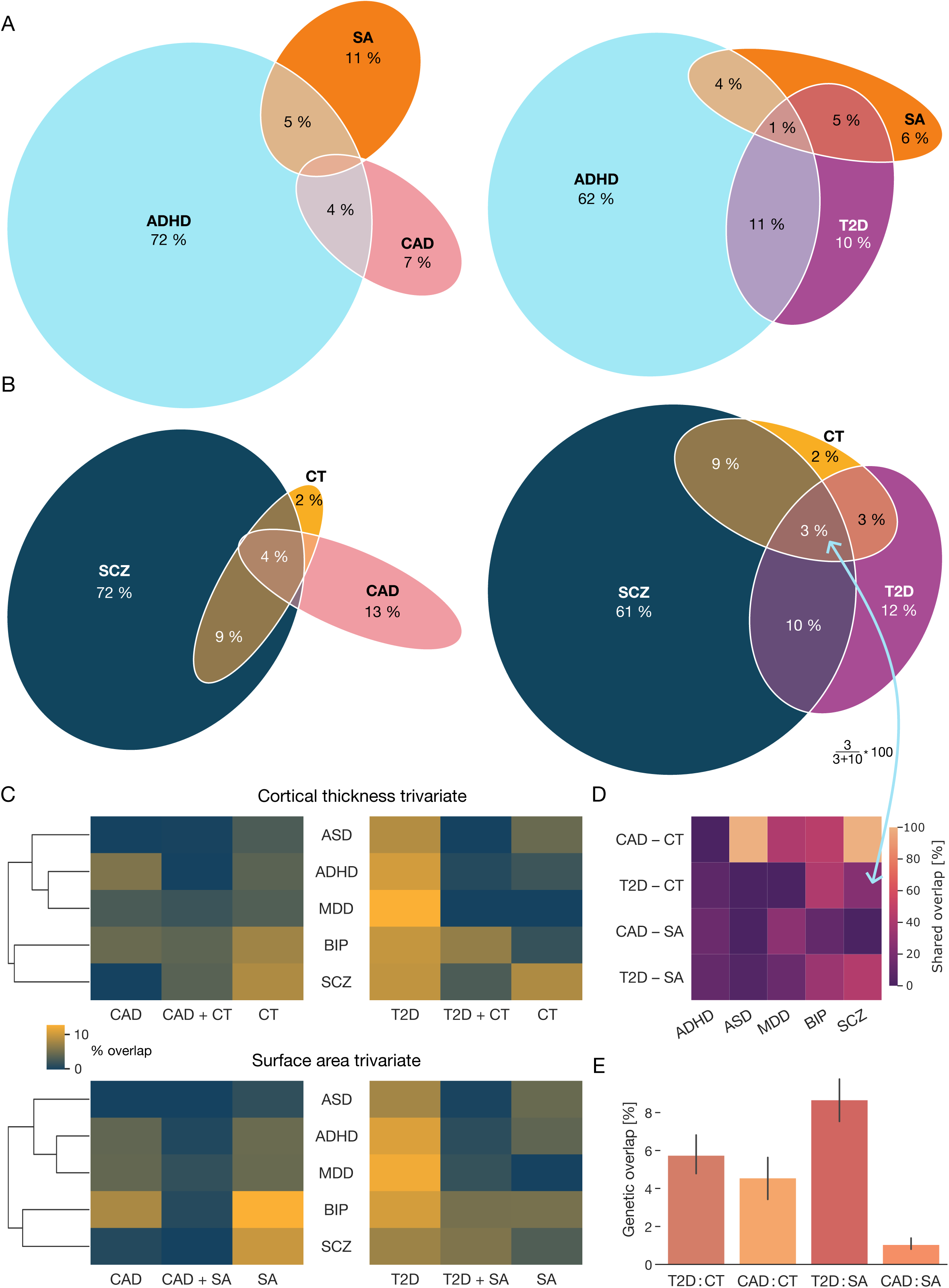
Trivariate overlap between psychiatric disorders, cardiometabolic disease and cortical morphology. **A.** Genetic overlap between ADHD, SA, and cardiometabolic disease. Based on trivariate MiXeR analyses, the Venn diagrams illustrate the extent of genetic overlap between cortical morphology (i.e., SA), ADHD, and cardiometabolic traits (CAD and T2D). Only genetic overlaps exceeding 1% are annotated. The complete set of overlaps and standard errors are provided in Sup. Fig. 4. **B.** Genetic overlap between SCZ, CT, and cardiometabolic disease. Genetic overlaps between CT, SCZ, and the cardiometabolic traits CAD and T2D are presented using Venn diagrams. **C.** Trivariate analyses for all five psychiatric disorders. The first three columns display overlaps between psychiatric disorders, cortical morphology, and CAD, while the second three columns display overlaps between psychiatric disorders, cortical morphology, and T2D. Results are first presented for CT followed by SA. Hierarchical clustering of disorders highlight their similarities in terms of morphology and cardiometabolic overlaps, shown separately for CT and SA. **D.** Zooming in on trivariate overlaps. The heatmap shows the percentage of genetic overlap between each psychiatric disorder and cardiometabolic disease (CAD or T2D) that is also shared with either CT or SA. Higher values indicate greater trivariate convergence across psychiatric, cardiometabolic, and cortical morphology domains. The calculation is demonstrated on SCZ, T2D, and CT, where the trivariate overlap accounted for 23% of the genetic overlap between SCZ and T2D. **E.** Genetic overlap between cortical morphology and cardiometabolic disease. The bar plot illustrates the average proportion of shared genetic variants between cortical morphology (CT and SA) and cardiometabolic traits (T2D and CAD) across all trivariate MiXeR analyses. Error bars indicate 95% confidence intervals. Collectively, the presented analyses revealed substantial genetic overlap between SCZ, CT, and T2D. In contrast, ADHD displayed a distinct genetic pattern, with smaller trivariate overlaps.

In contrast to ADHD, SCZ exhibited substantially larger trivariate overlaps with cortical morphology, and cardiometabolic disease. Given the stronger genetic overlap between SCZ and CT, we focused on presenting the overlaps between SCZ, CT, and cardiometabolic diseases (Fig. 2B). Results for SA are provided in Sup. Fig. 1. Notably, we did not observe trait-influencing variants uniquely shared between SCZ and CAD independent of CT. SCZ shared 9% of trait-influencing variants with CT, along with an additional 4% shared jointly among SCZ, CT, and CAD. The genetic overlap between SCZ and T2D (10%) was comparable to that between SCZ and CT (9%), with an additional 3% of trait-influencing variants shared among SCZ, T2D, and CT. In summary, our trivariate analyses revealed substantial genetic overlap among SCZ, CT, and cardiometabolic diseases, suggesting a shared genetic basis among neurodevelopmental and metabolic phenotypes. By comparison, ADHD exhibited a distinct genetic architecture characterized by smaller trivariate overlaps.

In order to investigate whether the observed genetic overlap is disease-specific, we extended our trivariate analysis to the additional psychiatric disorders (i.e., BIP, ASD, and MDD). The detailed results are provided in Sup. Fig. 2, 3. We then performed hierarchical clustering on the derived genetic overlaps with cardiometabolic disease separately for CT and SA, based on Ward distance (Fig. 2C). In other words, we searched among the five psychiatric disorders for similar patterns of overlap with cardiometabolic diseases and CT as well as SA. These post-hoc analyses again highlighted SCZ and ADHD as two disorders exhibiting distinct genetic overlap patterns. Furthermore, all five disorders demonstrated notable genetic overlap with T2D. Conversely, the degree of overlap with CAD and cortical morphology varied among disorders. SCZ and BIP exhibited the strongest genetic overlap with cortical morphology, while ADHD, BIP, and MDD showed the highest levels of genetic overlap with CAD. In contrast, ASD exhibited only minimal overlap with both CAD and cortical morphology.

Furthermore, we quantified how much of the genetic overlap between psychiatric disorders and cardiometabolic diseases is also shared with cortical morphology by computing the ratio of trivariate overlap to the total bivariate overlap for each disorder pair (i.e., trivariate / [trivariate + unique bivariate]) (Fig. 2D). We found that a substantial portion of the genetic overlap between psychiatric disorders and CAD was also shared with CT (ASD: 100%, SCZ: 100%, BIP: 46, MDD: 42%, ADHD: 0%).

Finally, we quantified the overall genetic overlap between cortical morphology and cardiometabolic disease by averaging trivariate MiXeR results across all psychiatric disorders. We observed a stronger average genetic overlap between T2D and both CT and SA compared to CAD (Fig. 2E). The overlap between CAD and SA was notably smaller, supporting a more prominent link between T2D and cortical morphology. Collectively, our results highlight potential cardiometabolic contributions to brain-related genetic risk for psychiatric disorders.

### Distinct causal relationships in psychiatric disorders

Our trivariate analyses demonstrated distinct genetic signatures of ADHD and SCZ. To follow up these results of genetic overlaps, we probed causal relationships between cortical morphology, psychiatric disorders and cardiometabolic diseases (Sup. Tab. 1). Focusing on the trait combinations with the highest genetic overlap, we further investigated the bidirectional causal relationships between ADHD, T2D, and SA, as well as SCZ, T2D, and CT, using inverse-variance weighted (IVW) Mendelian randomization (MR) (Fig. 3A).

**Figure 3:**
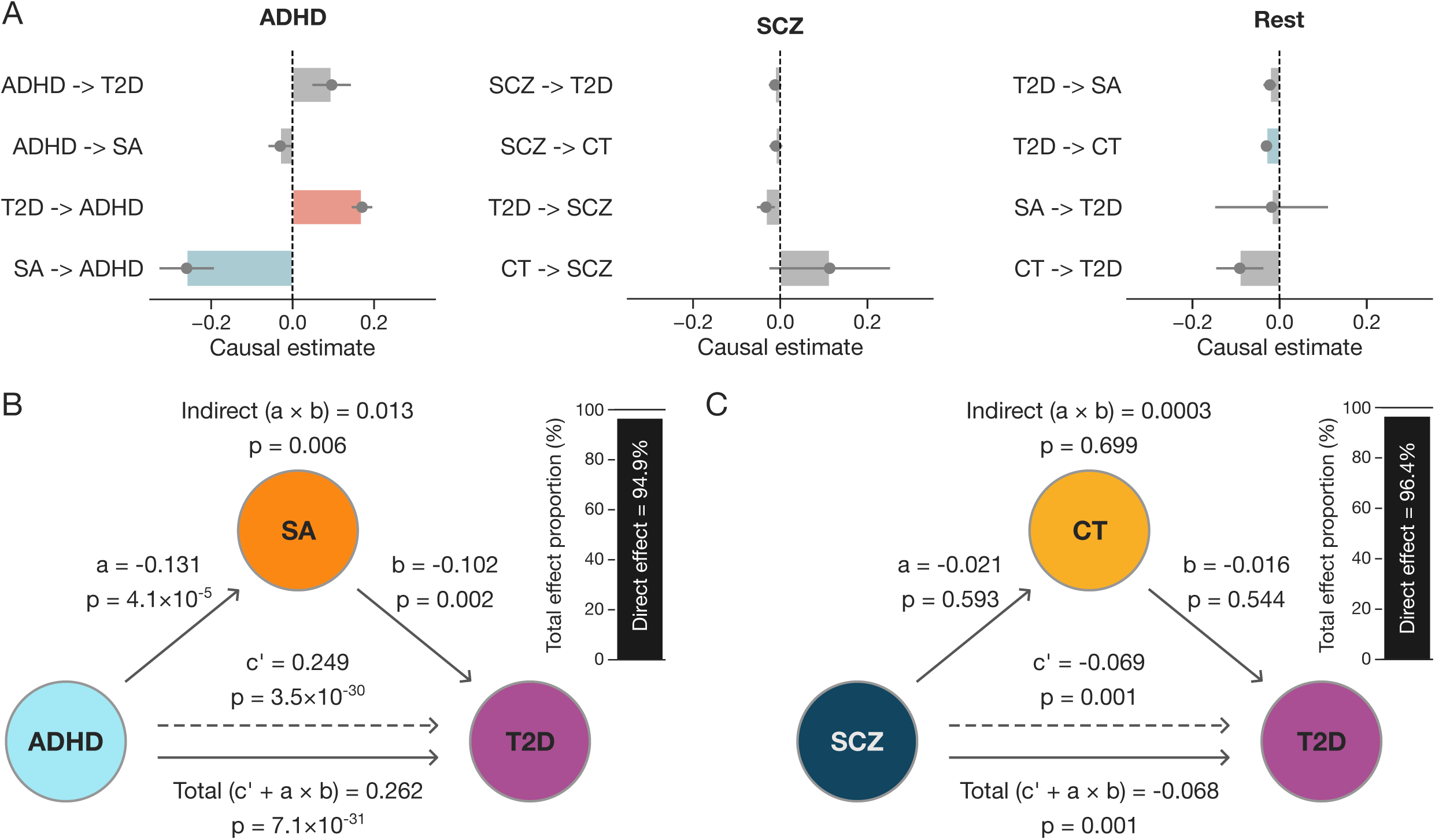
Causal and mediated genetic pathways from psychiatric traits to cardiometabolic risk. **A.** Bivariate causal effect mapping. Bar plots display the magnitude and direction of estimated causal effects from inverse-variance weighted (IVW) Mendelian randomization (MR) analyses focused on SCZ and ADHD. Based on prior findings, we conducted a series of bivariate IVW MR analyses to assess causal relationships among ADHD, T2D, and SA, followed by a second set of analyses among SCZ, T2D, and CT. Error bars represent standard errors of the causal effect estimates. Associations with FDR-corrected p-values > 0.05 are shown in grey. Full results are provided in Sup. Table 1. **B.** SA mediates the effect of ADHD on T2D. Mediation model evaluating surface area (SA) as a mediator of the association between ADHD (exposure) and T2D (outcome). Path coefficients (β) and corresponding p-values are shown for direct and indirect effects. The total effect (c′L+LaL×Lb) is reported below the model. A side bar indicates the proportion of the total effect attributable to the direct and indirect paths. **C.** CT does not mediate the effect of SCZ on T2D. Mediation model evaluating CT as a mediator between SCZ (exposure) and T2D (outcome), presented using the same structure as panel **B**. Presented results suggest that SA may partially mediate the genetic link between ADHD and T2D, whereas the association between SCZ and T2D likely reflects independent or parallel genetic pathways.

We found evidence that genetic liability for T2D has a causal effect on ADHD risk, with higher T2D liability associated with increased genetic susceptibility to ADHD (β = 0.13, FDR-correct *p*_FDR_ = 9.5×10^-11^). In addition, lower SA was associated with increased genetic risk for ADHD (β = –0.26, *p*_FDR_ = 0.0003).

In contrast to the findings for ADHD, IVW MR did not reveal any significant causal effect of SCZ on CT or T2D. Finally, we identified a significant negative effect of T2D on CT (β = –0.03, *p*_FDR_ = 0.03), suggesting that increased genetic liability for T2D may contribute to cortical thinning. These findings suggest that ADHD is embedded in a bidirectional causal network involving SA and T2D risk, whereas SCZ may be indirectly shaped by T2D liability through its effects on CT.

Building on our bivariate MR results, we next examined whether cortical morphology mediates the associations between psychiatric disorders and cardiometabolic disease. Using GenomicSEM-based mediation models, we focused on the strongest causal signals identified earlier: ADHD–SA–T2D and SCZ–CT–T2D (Fig. 3B). In the ADHD model, we observed a significant indirect effect through SA on the association between ADHD and T2D (β = 0.013, *p* = 0.006), indicating partial mediation. This was supported by significant paths from ADHD to SA (β = –0.131, *p* = 4.1×10^-5^) and from SA to T2D (β = –0.102, *p* = 0.002). The direct effect of ADHD on T2D remained strong (β = 0.249, *p* = 3.5×10^-30^), with SA accounting for approximately 5% of the total effect.

In contrast, CT did not mediate the association between SCZ and T2D (Fig. 3B). Neither the effect of SCZ on CT (β = –0.021, *p* = 0.593) nor the effect of CT on T2D (β = – 0.016, *p* = 0.544) was significant, resulting in a negligible indirect effect (β = 0.0003, *p* = 0.69). The direct effect of SCZ on T2D (β = –0.069, *p* = 0.001) accounted for nearly the entire total effect. These findings suggest that while SA may play a modest intermediary role in the genetic link between ADHD and cardiometabolic risk, the association between SCZ and T2D likely reflects independent or parallel genetic pathways, rather than mediation through brain structure.

### Shared loci underlying psychiatric and cardiometabolic comorbidity

To gain deeper biological insights into the mechanisms underlying multimorbidity, we aimed to identify shared biological pathways linking mental health with cardiometabolic disease and cortical morphology. To do so, we began with a conjunctional FDR analysis (conjFDR) that identified genetic variants that are shared between ADHD and T2D, ADHD and SA, SCZ and T2D, as well as SCZ and CT.

The first conjFDR analysis, conducted at a threshold of conjFDR < 0.05, revealed 93 lead SNPs jointly associated with ADHD and T2D (Fig. 4A). Applying the same threshold of conjFDR < 0.05, we identified 17 lead SNPs jointly associated with ADHD and T2D (Fig. 4B). We then targeted loci shared between SCZ and T2D. The conjFDR analysis revealed 320 lead SNPs jointly associated with SCZ and T2D (Fig. 4C). Finally, we identified 38 lead SNPs jointly associated with SCZ and CT (Fig. 4D). Results using a stricter conjFDR threshold of <0.01 are presented in Sup. Fig. 4.

**Figure 4:**
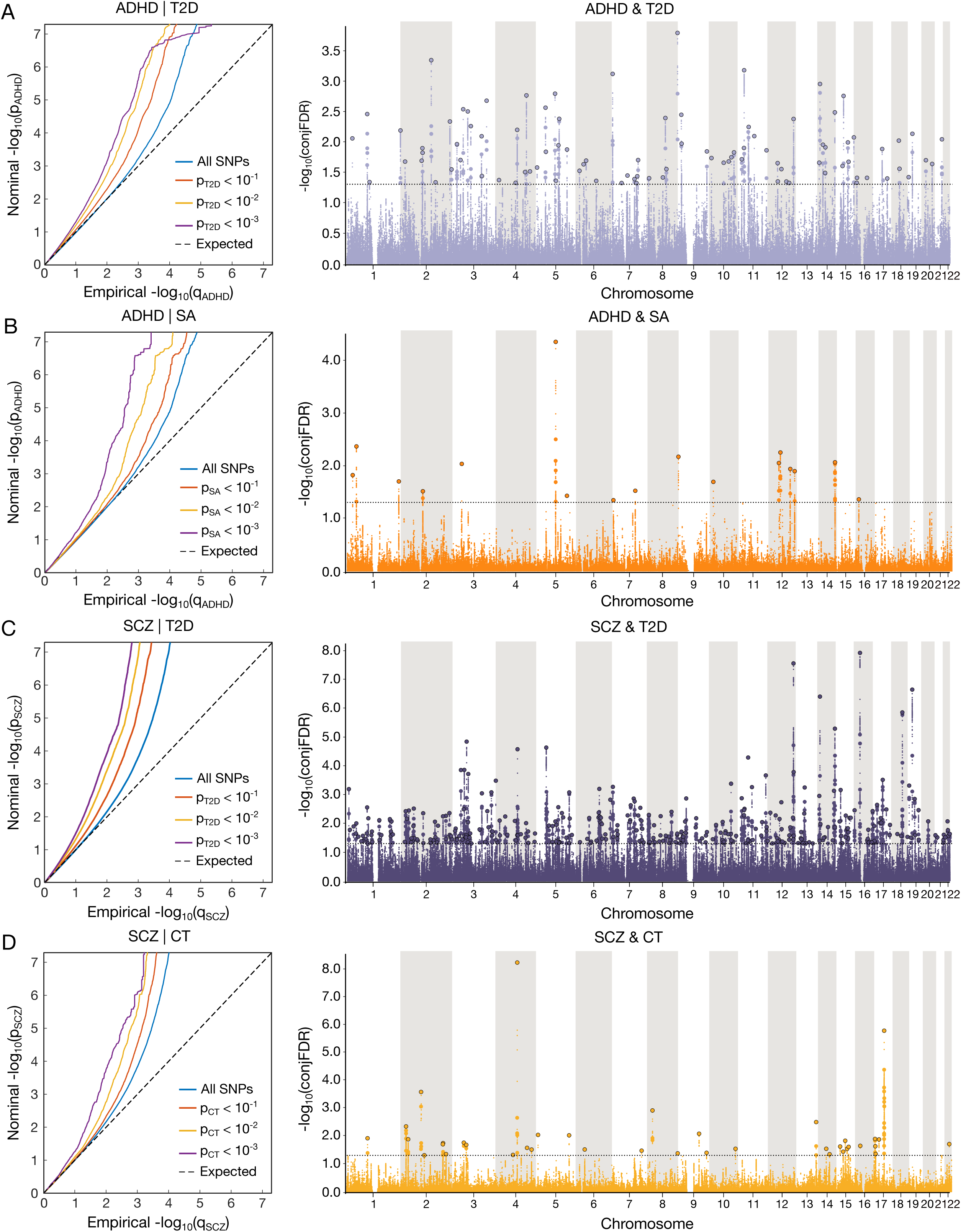
Cross-trait genetic enrichment and shared loci. We leverage the presented genetic overlaps to identify shared variants between cardiometabolic disease, psychiatric disorders, and cortical morphology. Specifically, we search for SNPs jointly associated between ADHD and SA (**A**), ADHD and T2D (**B**), SCZ and T2D (**C**), as well as SCZ and CT **(D).** For these four analyses, the Q-Q plot first compares observed (y-axis) and expected p-values (x-axis) to evaluate whether there is sufficient cross-trait enrichment of genetic associations, which is a prerequisite for conducting the conjFDR analysis. The diagonal line represents the expected distribution under the null hypothesis. The colored lines represent Q-Q plots for SNP subsets filtered by their association strength with cardiometabolic disease. Points above the diagonal indicate enrichment, where observed p-values are stronger (smaller) than expected, suggesting associations between traits or enrichment in specific regions. The associated Manhattan plot displays genetic variants shared by the respective two traits. Each Manhattan plot shows the –log_10_ transformed conjFDR values for each SNP on the y-axis. SNPs with conjFDR < 0.05 (i.e., −log_10_ conjFDR > 1.3) are shown with enlarged data points. A black circle around the enlarged data points indicates the most significant SNP in each linkage disequilibrium block. The large number of shared SNPs across conjFDR analyses provides evidence for shared genetic pathways underlying multimorbidity between psychiatric and cardiometabolic disorders.

Following the identification of shared SNPs, we mapped all candidate variants to genes using OpenTargets (see Methods). The shared candidate SNPs between ADHD and T2D were mapped to 42 genes, between ADHD and SA to 15 genes, between SCZ and T2D to 213 genes, and SNPs shared between SCZ and CT were mapped to 32 genes (Sup. Tab. 2). In summary, conjFDR analyses followed by gene mapping identified extensive cross-trait genetic overlap, revealing numerous genes jointly associated with both cardiometabolic and psychiatric disorders.

### Overlapping biological pathways of multimorbidity

In the next step, the four gene sets derived from the combination of conjFDR results and gene mapping were sequentially tested for enrichment of biological pathways from the Kyoto Encyclopedia of Genes and Genomes (KEGG) and Reactome using DAVID (Database for Annotation, Visualization, and Integrated Discovery). KEGG and Reactome are comprehensive databases that provide insights into biological processes, molecular interactions, and disease mechanisms. We calculated the fold enrichment of each KEGG and Reactome pathway for each conjFDR analysis, providing a measure of overrepresentation of pathways in the identified gene sets (Sup. Tab. 3).

Analysis of genes jointly associated with ADHD and SA revealed enrichment in multiple biological pathways (Fig. 5A). These could be broadly grouped into vascular signaling processes (e.g., VEGFR2-mediated vascular permeability, VEGFA–VEGFR2 signaling), metabolic and endocrine regulation (e.g., TP53-regulated metabolic genes, adipocytokine signaling, transcriptional regulation of MITF-M), and cell signaling mechanisms (e.g., RAB GEFs exchange). Additional enrichment was observed in pathways related to cancer biology (e.g., endometrial cancer) and longevity regulation across species.

**Figure 5:**
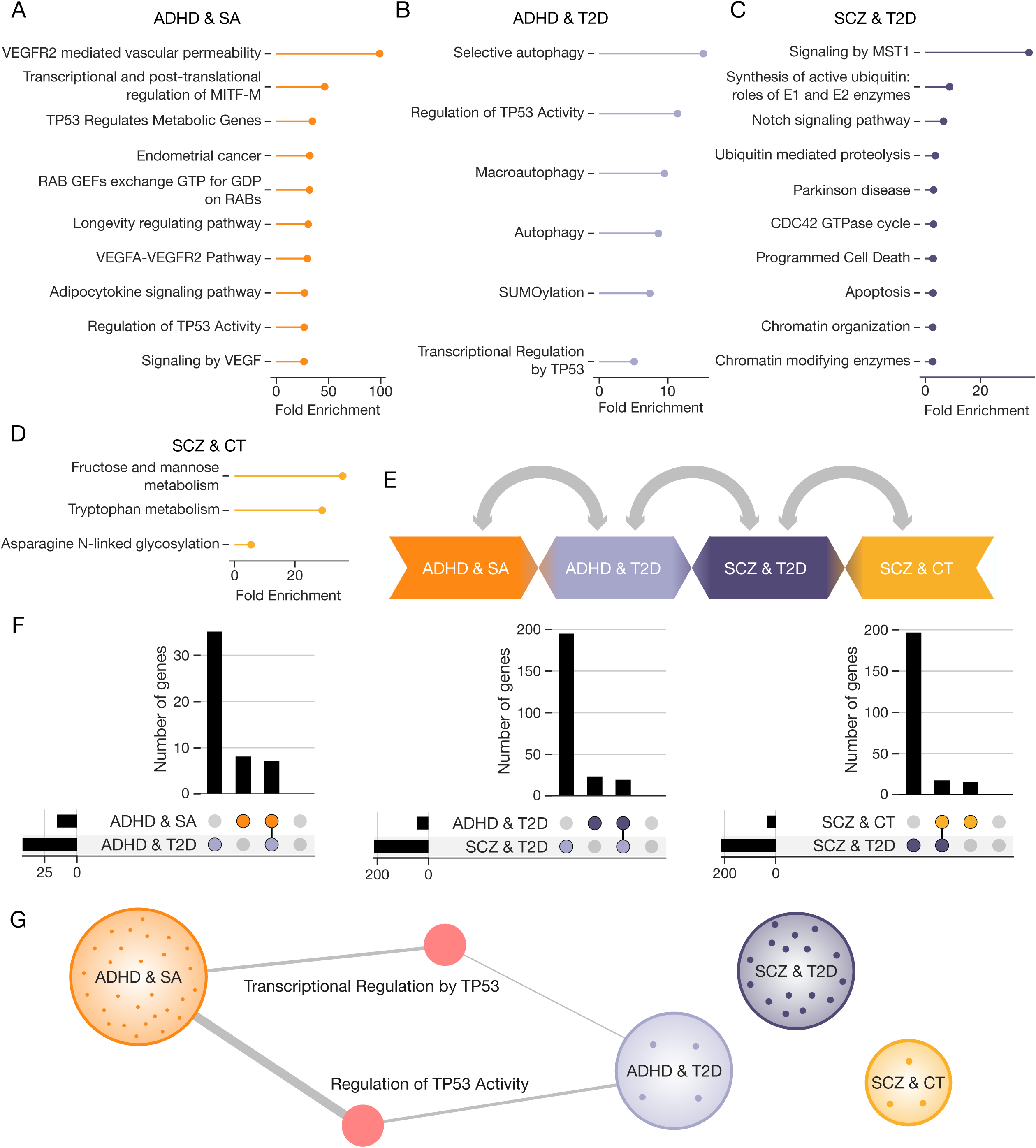
Biological pathways underlying multimorbidity. Genes mapped using FUMA from the four conjFDR analyses were assessed for pathway enrichment using the DAVID tool, focusing on KEGG and Reactome databases. The bar plots illustrate the top enriched biological pathways (up to ten per analysis) for genes associated with each of the four conjFDR results: genes mapped to SNPs jointly associated with ADHD and SA **(A)**, ADHD and T2D **(B)**, SCZ and T2D **(C)**, and SCZ and CT **(D)**. **E**. Comparison of shared genes and pathways. Each tile in the diagram represents a conjFDR analysis, and we examined the overlap in shared genes and enriched pathways between neighboring analyses. **F.** Gene overlaps visualized using an UpSet plot. The horizontal bar chart on the left displays the total number of identified genes for each conjFDR analysis. The intersection matrix at the center represents overlaps between the analyses, where colored dots indicate set membership and connected lines between dots denote intersections of multiple gene sets. The vertical bar chart above the matrix shows the size of each intersection. **G.** Overlap in biological pathways. The network diagram highlights the shared biological pathways across the gene sets. The four large nodes represent the four conjFDR analyses, with dots inside each node indicating biological pathways unique to that analysis. The thickness of the edges connecting pathways corresponds to their respective fold enrichment, illustrating the strength of the associations. Pathway enrichment analyses suggested that *TP53*-related transcriptional regulation may represent a potential point of convergence between brain structure, psychiatric disorders, and cardiometabolic traits.

Pathway enrichment analysis of genes shared between ADHD and T2D highlighted several pathways related to autophagy and intracellular degradation, including selective autophagy, macroautophagy, and general autophagy reflecting processes involved in cellular quality control and metabolic balance (Fig. 5B). Enrichment was also observed in post-translational modification, specifically the SUMOylation pathway, as well as in TP53-related transcriptional and stress-response pathways, including Regulation of TP53 Activity and Transcriptional Regulation by TP53.

For genes jointly associated with SCZ and T2D, enriched pathways reflected cell death and survival processes (programmed cell death, apoptosis, signaling by MST1), protein homeostasis (ubiquitin-mediated proteolysis, synthesis of active ubiquitin via E1 and E2 enzymes), and chromatin regulation (chromatin-modifying enzymes, chromatin organization) (Fig. 5C). Additional pathways involved developmental and intracellular signaling, including the Notch signaling pathway and the CDC42 GTPase cycle.

Finally, pathway enrichment analysis of genes shared between SCZ and cortical thickness revealed involvement of core metabolic pathways, including fructose and mannose metabolism, tryptophan metabolism, and protein modification (asparagine N-linked glycosylation) (Fig. 5D).

As a final step in exploring potential points of convergence across cortical morphology, psychiatric disorders, and metabolic disease, we examined overlapping genes and enriched pathways between selected pairs of conjFDR results (Fig.L5E). Specifically, we performed within-disorder comparisons (ADHD & SA vs. ADHD & T2D; SCZ & CT vs. SCZ & T2D) to assess whether genetic links to brain and metabolic traits arise through shared mechanisms, as well as a cross-disorder comparison (ADHD & T2D vs. SCZ & T2D) to determine whether distinct psychiatric conditions share common pathways to cardiometabolic risk. These comparisons revealed eight shared genes in the within-disorder analyses for both ADHD (ADHD & SA vs. ADHD & T2D) and SCZ (SCZ & CT vs. SCZ & T2D), and seven shared genes in the cross-disorder comparison of ADHD & T2D and SCZ & T2D. This overlap points to possible shared biological mechanisms linking brain structure, psychiatric liability, and cardiometabolic risk (Fig.L5F).

Notably, only two biological pathways were shared across any of the comparisons: Regulation of TP53 Activity and Transcriptional Regulation by TP53, both linking ADHDL&LSA and ADHDL&LT2D (Fig.L5G). This convergence highlights a potential role for TP53-related transcriptional regulation in bridging genetic risk for ADHD across brain structure and metabolic dysfunction.

## Discussion

In this study, we systematically investigated the shared and distinct genetic architecture linking psychiatric disorders, cardiometabolic diseases, and cortical morphology. Using a combination of trivariate MiXeR, MR, and pathway enrichment analyses, we provide converging evidence that ADHD and SCZ exhibit overlapping yet distinct patterns of genetic architecture. SCZ showed substantial polygenic overlap with both CT and T2D, suggesting a shared genetic basis among neurodevelopmental and metabolic phenotypes. In contrast, ADHD displayed a more distinct genetic profile, with less polygenic overlap with cortical morphology but stronger connections to cardiometabolic traits. Overall, our results suggest that multimorbidity between psychiatric and cardiometabolic conditions is shaped by a combination of disorder-specific mechanisms and partially shared biological pathways, where common regulatory processes may intersect with distinct trait-specific genetic architectures.

Despite widespread recognition of psychiatric–cardiometabolic comorbidity, the extent to which this overlap is driven by shared genetic factors or mediated through brain structure remains unclear. Using trivariate genetic overlap analysis, we observed substantial genetic overlap among SCZ, CT, and T2D. However, mediation analysis provided no evidence for CT as a pathway linking SCZ to cardiometabolic risk. On the other hand, ADHD showed a smaller trivariate overlap with SA and T2D, yet mediation analysis revealed that SA partially accounted for the genetic association between ADHD and T2D. Notably, TP53- related pathways were enriched in both ADHD & SA and ADHD & T2D overlaps, pointing to transcriptional stress regulation as a potential shared mechanism linking neurodevelopment and metabolic vulnerability in ADHD. As such, SCZ may involve broader systemic and neurodevelopmental mechanisms, whereas ADHD appears to follow more specific genetic pathways linking it to cardiometabolic risk, potentially mediated by cortical surface area.

These findings prompt the examination of biological mechanisms that might explain the shared genetic architecture between brain structure, psychiatric conditions, and cardiometabolic diseases. Brain architecture has long been recognized as a fundamental determinant of cognition, mental resilience, and susceptibility to psychiatric disorders^47–49^. Despite overlapping symptomatology, psychiatric disorders often exhibit distinct patterns of subcortical and cortical alterations^50^. As an example, SCZ is primarily associated with widespread reductions in CT, with particularly pronounced effects in frontal and temporal regions^34^. These effects on CT are notably stronger than those observed for SA. In contrast, ADHD is more strongly linked to reduced SA, especially in the frontal, cingulate, and temporal regions, while differences in CT tend to be subtle in childhood and largely absent in adolescence and adulthood^51^.

At least partially, these differences can be explained by the differences in genetic architecture. Here, we observed that SCZ aligned more closely with cortical morphology compared to ADHD. In line with our results, a significantly higher number of SNPs have been previously identified as jointly associated with SCZ and CT compared to ADHD and CT^52^. Almost 50% of genes linked to SCZ were previously associated with brain morphology^53^. However, the genetic correlations between SCZ and cortical morphology were close to zero, consistent with previous findings^40^. Conversely, ADHD showed a significant genetic correlation with SA^40^. This link is further supported by our MR results, which revealed a directional effect of SA on ADHD as well as a significant mediation pathway linking ADHD to T2D through SA. Although our MR analysis did not identify a significant effect of SCZ on global CT, previous studies have reported such associations when focusing on regional cortical thickness measures^54^. Collectively, these findings suggest that although ADHD shares fewer variants with SA than SCZ does with CT, the effects of these variants may be more directionally consistent, enabling both causal inference and mediation.

Pathway enrichment analysis highlighted a stronger metabolic signature in the genetic overlap between ADHD and cortical morphology than in the overlap with SCZ. For ADHD & SA, we observed enrichment in several metabolic-related pathways suggesting that metabolic regulation may be a key biological mechanism linking neurodevelopmental alterations in cortical SA with ADHD liability. In contrast, the SCZ & CT overlap yielded fewer enriched pathways, with tryptophan metabolism standing out as a plausible contributor. Tryptophan metabolism, which is influenced by diet, inflammation, and insulin resistance, may represent a more indirect or environmentally sensitive mechanism linking metabolic dysfunction to cortical morphology in SCZ^55^. These findings provide a foundation for exploring how cardiometabolic traits might shape brain structure in ways that contribute to psychiatric vulnerability.

A large body of research documents the tight connection between cardiometabolic traits and brain architecture^56^. Specifically, obesity, hypertension, high cholesterol, diabetes, as well smoking have been linked to brain atrophy^42,57,58^. In addition, MR studies indicate that higher BMI may causally contribute to reduced gray matter volume^59^. Our results further show that T2D exerts a modest but significant causal effect on cortical thickness. These adverse effects on brain tissue and gray matter were further reported to increase in severity with the number of chronic diseases, particularly when metabolic and cardiometabolic conditions are involved^60^. Going beyond cross-sectional evidence, individuals with multimorbidity also show a steeper decline in total brain tissue volume and accelerated ventricular enlargement over time compared to their non-multimorbid peers^61^. Consequently, adults with T2D might exhibit cognitive dysfunction, characterized by impairments in attention, psychomotor speed, planning, executive functions, and memory^62^. Although the clinical and imaging evidence linking metabolic health to brain structure is well established, the extent to which these associations reflect shared genetic architecture or directional causal effects remains poorly understood.

To address this uncertainty, researchers have begun applying genomic approaches to disentangle common heredity from causation in cardiometabolic–brain connections. Genetic correlation studies have revealed significant overlapping heritability between adiposity-related traits (e.g. BMI) and brain structure measures^63,64^, suggesting that some of the same genetic variants may predispose individuals to both metabolic dysfunction and neuroanatomical changes. Here, we observed substantial polygenic overlap among T2D and CT as well as SA supporting the existence of shared genetic components across these domains. Our findings are consistent with recent metabolomic studies showing that circulating metabolic markers, many of which are altered in T2D^65^, are genetically associated with cortical morphology^56^. The collective evidence thus points to a shared genetic basis between metabolic health and brain structure, raising the question of how these interconnected pathways may also contribute to psychiatric disorders and their cardiometabolic comorbidities.

Psychiatric and cardiometabolic disorders are known to co-occur at high rates, prompting an investigation into their shared genetic underpinnings. We observed that the genetic relationships between psychiatric and cardiometabolic disorders are heterogeneous, suggesting disorder-specific patterns of comorbidity. Among the five disorders, ADHD and MDD showed the strongest genetic correlations and the most prominent overlap with CAD, in contrast to the weaker associations observed for SCZ and ASD. The genetic architecture of MDD was already shown to be more similar to cardiometabolic diseases than those of SCZ and BIP based on patterns of genetic overlap with metabolic markers^66^. This partly shared genetic architecture between CAD and MDD as well as ADHD might be one of the reasons why genetic liability for ADHD was proposed as a potential causal factor for CAD^67^. In another study, childhood ADHD problems were predictive of multiple cardiovascular risk factors by mid-life^68^. However, ADHD comorbidity with cardiometabolic disorders was proposed to be mainly driven by environmental factors rather than genetics^32^. Therefore, while genetic overlap may indicate shared biological pathways, environmental and lifestyle factors likely play a crucial role in determining whether this genetic risk translates into actual disease manifestation.

In comparison to CAD, all studied psychiatric disorders displayed substantial overlap with T2D based on trivariate MiXeR analyses. This aligns with prior evidence showing high comorbidity between psychiatric disorders and T2D, with suggestions of shared genetic underpinnings^32,69^. These genetic links between T2D and psychiatric disorders may be particularly pronounced in neurodevelopmental conditions^70^. Notably, despite a near-zero genetic correlation, SCZ and T2D have been shown to share substantial polygenic overlap^29^. Moreover, genes jointly associated with SCZ and T2D have been found to show high expression in brain tissue and immune cells, and are enriched for pathways related to neurodevelopment, synaptic signaling, immune function, intracellular transport, and metabolic regulation^29,70^. This brain- and immune-enriched functional profile aligns with our findings, suggesting that the SCZ & T2D genetic overlap may involve shared neurodevelopmental and systemic biological pathways. Collectively, our findings support distinct genetic architectures underlying psychiatric–cardiometabolic comorbidity. The SCZ & T2D overlap may involve brain-related mechanisms and indirect pathways, such as antipsychotic medication effects or lifestyle changes. In contrast, ADHD and MDD appear to have a more direct genetic predisposition to cardiometabolic conditions, likely compounded by behavioral and environmental risk factors.

Pathway enrichment analyses also suggested distinct biological mechanisms underlying the genetic overlap between T2D and psychiatric disorders. For ADHD and T2D, we observed strong enrichment in autophagy-related and metabolic stress pathways, including selective autophagy, SUMOylation, and TP53 regulation. These results point to systemic metabolic processes as a shared mechanism, which aligns with prior evidence linking ADHD to insulin resistance, oxidative stress, and energy metabolism abnormalities^71,72^. Overall, the enrichment profile suggests that metabolic vulnerability may represent key biological links between ADHD and T2D. In contrast, the SCZ & T2D overlap was enriched for pathways involved in protein homeostasis, chromatin organization, and immune regulation. These processes, including disruptions in protein turnover, epigenetic control, and neuroimmune signaling, may underlie both systemic and neurodevelopmental dysregulation in SCZ^29^. Taken together with the distinct enrichment profile observed for ADHD, our findings support the notion that ADHD and SCZ may follow different biological routes to cardiometabolic comorbidity.

In conclusion, we demonstrated how studying trivariate genetic overlaps can provide novel and more refined insights into the genetic etiology of multimorbidity. In doing so, we uncovered patterns of genetic overlap among psychiatric, cardiometabolic, and brain structural traits that are not detectable using traditional bivariate methods. Our findings highlight a substantial polygenic overlap between SCZ, T2D, and CT, suggesting that systemic processes such as neuroimmune signaling and cellular maintenance may contribute to their shared liability. In contrast, ADHD showed stronger genetic ties to SA and T2D, with enrichment in metabolic stress and autophagy-related pathways, pointing to a more direct, metabolically driven route to comorbidity. These disorder-specific signatures emphasize the importance of integrative, biologically informed approaches to understanding psychiatric–cardiometabolic comorbidity and may ultimately inform more targeted strategies for prevention and treatment.

## Methods

### Participant cohorts

This study sets out to disentangle genetic overlaps between mental disorders, cardiometabolic disease, and brain organization. The study of genetic overlap is based on genetic architecture represented by genome-wide association studies (GWAS) (Table 1). The GWAS schizophrenia was retrieved from the Psychiatric Genomics Consortium (PGC) and included 53,386 participants diagnosed with SCZ and 77,258 controls^73^. Similarly, GWAS of ADHD was retrieved from PGC and consisted of 38,691 participants with ADHD diagnosis and 186,843 controls^74^. The summary statistics of other psychiatric disorders are summarized in Table 1. The genetic architecture of T2D was based on data from 1,569,731 controls and 242,283 cases^75^. CAD was represented by GWAS from the study of 181,522 participants with CAD and 984,168 controls^76^. Finally, GWAS of CT were based on the UK Biobank (UKB) sample. We accessed UKB data under accession number 27412. The design, participant composition, and data collection protocols of the UKB have been described in detail elsewhere^77^. We selected 39,098 unrelated individuals (KING cutoff = 0.0884)^78^ with T1 MRI data. All GWAS summary statistics are based on Human Genome Build 37 (GRCh37/hg19).

To minimize the confounding effects of ancestral differences in linkage disequilibrium (LD) structure and due to the limited availability of sufficiently powered multi-ancestry samples, we restricted our analysis to individuals of European ancestry. We excluded variants within the extended major histocompatibility complex (MHC) region (chr6: 25–35 Mb) for all subsequent analyses. We further excluded the 8p23 inversion region (chr8: 7.2– 12.5 Mb) and APOE locus (chr19:42–47 Mb) due to their high LD and complex structure. The local ethics committees approved all GWASs used in the current study, and all participants provided informed consent. Regional Committees for Medical Research Ethics - South-East Norway has evaluated the current protocol and found that no additional institutional review board approval was necessary because no individual data were used.

### Statistical analyses

#### Calculating genetic correlation

We applied cross-trait LDSC^79^ to estimate genetic correlations between psychiatric disorders and cardiometabolic diseases. In addition, we used univariate LDSC to estimate SNP-based heritability for each trait. GWAS summary statistics were processed following recommended guidelines, including standard “munging” procedures. Precalculated LD scores from the European 1000 Genomes reference cohort were used (https://data.broadinstitute.org/alkesgroup/LDSCORE/eur_w_ld_chr.tar.bz2). P-values were corrected for multiple testing using the Benjamini–Yekutieli procedure for controlling the false discovery rate^80^.

#### Quantifying trivariate genetic overlap

To quantify the genetic overlap between psychiatric disorders, cardiometabolic disease and cortical morphology, we leveraged the trivariate MiXeR tool^46^. The trivariate MiXeR model uses summary statistics from GWAS to quantify the polygenic overlap between three complex phenotypes beyond global genetic correlation. While genetic correlation is a popular approach for quantifying genetic overlap, it is restricted to detecting overlaps in pairs of phenotypes where most variants have effects in similar or opposing directions^81^.

The trivariate MiXeR extends the widely used univariate MiXeR^82^ that calculates the number of genetic variants influencing a phenotype (i.e., polygenicity) and bivariate MiXeR model^83^ that quantifies the overlapping polygenic components between two phenotypes regardless of the effect directions. In our study, we also used univariate MiXeR to estimate the polygenicity of each phenotype individually prior to trivariate modeling. Despite the important insights that studying pairwise genetic overlaps brought^12,14^, genetic correlation and bivariate MiXeR cannot directly estimate genetic overlap across three phenotypes.

Similarly to previous MiXeR implementations, the trivariate MiXeR decomposes the observed genetic effects into shared causal variants affecting multiple traits and unique causal variants affecting only one trait. By iteratively adjusting the model parameters, this tool identifies the configuration (number and size of shared/unique effects) that best fits the observed data, maximizing the likelihood of the summary statistics under the model. Therefore, we used this tool to estimate the total number of shared and unique trait-influencing variants among psychiatric disorders, cardiometabolic disease and cortical morphology. Results presented in the main body of the article represent optimal estimates across 100 independent optimization runs, while results for each of the 100 runs are available in the Sup. Fig. 5.

#### Estimating causal relationships

We first conducted bidirectional Mendelian Randomization (MR) analyses to investigate potential causal relationships between pairs of psychiatric disorders, cortical morphology, and cardiometabolic diseases. MR was performed using the TwoSampleMR R package (v0.6.15) applied to GWAS summary statistics. Instrumental variables were selected as genome-wide significant SNPs (p < 5×10^⁻□^), and clumping was performed using PLINK with the following parameters: clump_p = 1, clump_r2 = 0.001, and clump_kb = 10000, using the 1000 Genomes Phase 3 European reference panel (n = 503). All other settings were kept at default. Causal estimates were obtained using the Inverse Variance Weighted (IVW) method. All estimates with associated standard errors and p-values are available in Sup. Tab. 1. We also include estimates using weighted median^84^, MR-Egger^85^, Simple Mode, and Weighted Mode^86^ methods (Sup. Tab. 1).

To determine whether brain morphology mediates the relationship between psychiatric and cardiometabolic traits, we conducted mediation analysis using the GenomicSEM R package (v0.0.5). We specified trivariate models with the psychiatric trait as the exposure, the cardiometabolic disease as the outcome, and cortical morphology as the mediator. SNP effect estimates and standard errors were extracted from GWAS summary statistics for each phenotype. We calculated direct, indirect, and total effects, and evaluated the significance of each path.

#### Mapping shared genetic loci

After the explorations of genetic overlap between multiple complex traits, we focused on the identification of shared SNPs among distinct traits. Specifically, ConjFDR can detect loci jointly associated with two phenotypes (e.g., SCZ and T2D)^81^. ConjFDR is an extension of the condFDR tool estimating the probability that a genetic variant is a false positive for one trait, given its association with the other traits^81^. In other words, this method adjusts the ranking of test statistics for a primary phenotype by incorporating information from the association of the same genetic variants with a secondary phenotype. This re-ranking increases the power to detect genetic signals shared between phenotypes by leveraging pleiotropic effects. After calculating the condFDR for each trait pair, the resulting conjFDR is defined as the maximum of condFDR values across all traits being analyzed^81,87^. P-values are adjusted for inflation using a genomic inflation control method^81,87^. Consistent with previously published studies, we used thresholds of 0.01 and 0.05 for adjusted p-values^29,31,88^.

#### Variant-to-gene mapping via functional annotation

We further analyzed genetic variants identified using conjFDR analysis at a threshold of conjFDR < 0.05. To reduce redundancy due to LD, we performed clumping on these SNPs using an LD threshold of r² < 0.6, resulting in a set of independent index variants. For each index SNP, we then identified all SNPs in high LD (r² ≥ 0.6) based on the 1000 Genomes Project European reference panel, defining them as candidate SNPs. These candidate variants were mapped to genes using the Variant-to-Gene (V2G) pipeline from Open Targets Genetics^89^, which integrates evidence from expression and protein QTLs, chromatin conformation (e.g., promoter–enhancer interactions), in silico functional predictions, and genomic proximity to transcription start sites. We retained only gene assignments with a V2G score greater than 0.3, indicating moderate-to-strong functional support.

#### Pathway enrichment analysis

To gain insight into the biological mechanisms underlying the identified loci, we performed pathway enrichment analysis on the gene sets derived from each conjFDR analysis. Specifically, we used the Database for Annotation, Visualization, and Integrated Discovery (DAVID) to map the genes identified by OpenTargets to curated biological pathways^90,91^. We focused on two major pathway databases: the Kyoto Encyclopedia of Genes and Genomes (KEGG)^92^ and Reactome^93^, which provide comprehensive annotations of molecular functions, biological processes, and disease-related pathways. For each of the four gene sets, enrichment was assessed separately using DAVID’s functional annotation tool. We calculated fold enrichment for each KEGG and Reactome pathway to determine the degree to which each pathway was overrepresented relative to what would be expected by chance. The resulting enrichment values provide an indication of the biological relevance of the gene sets to specific pathways.

## Data Availability

All GWAS summary statistics used in this study are publicly available through their respective consortia or original publications, as detailed in Table 1. Tools used for genetic overlap, causal inference, and functional annotation are also publicly available:

- cleansumstats (GWAS preprocessing): https://github.com/BioPsyk/cleansumstats
- LDSC: https://github.com/bulik/ldsc, https://github.com/comorment/ldsc
- Trivariate MiXeR: https://github.com/precimed/mix3r
- TwoSampleMR: https://mrcieu.github.io/TwoSampleMR
- GenomicSEM: https://github.com/GenomicSEM/GenomicSEM
- conjFDR: https://github.com/precimed/pleiofdr
- Open Targets Genetics (V2G mapping): https://genetics.opentargets.org/
- DAVID (pathway enrichment): https://davidbioinformatics.nih.gov/

All processed data and intermediate results (e.g., trivariate MiXeR outputs, mediation estimates, gene-pathway mappings) are available from the corresponding author upon reasonable request.

## Acknowledgements

This work was partly performed on the TSD (Tjeneste for Sensitive Data) facilities, owned by the University of Oslo, operated and developed by the TSD service group at the University of Oslo, IT-Department (USIT). Computations were also performed on resources provided by UNINETT Sigma2 - the National Infrastructure for High-Performance Computing and Data Storage in Norway (NS9666S).

## References

1. Felsky, D., Cannitelli, A. & Pipitone, J. Whole Person Modeling: a transdisciplinary approach to mental health research. Discov. Ment. Health 3, 16 (2023).

2. Kendler, K. S. Are Psychiatric Disorders Brain Diseases?—A New Look at an Old Question. JAMA Psychiatry 81, 325–326 (2024).

3. Bennett, F. C. & Molofsky, A. V. The immune system and psychiatric disease: a basic science perspective. Clin. Exp. Immunol. 197, 294 (2019).

4. Ohrnberger, J., Fichera, E. & Sutton, M. The relationship between physical and mental health: A mediation analysis. Soc. Sci. Med. 195, 42–49 (2017).

5. Casey, D. E. Metabolic issues and cardiovascular disease in patients with psychiatric disorders. Am. J. Med. Suppl. 118, 15–22 (2005).

6. Goldfarb, M. et al. Severe Mental Illness and Cardiovascular Disease: JACC State-of-the-Art Review. J. Am. Coll. Cardiol. 80, 918–933 (2022).

7. Mitchell, K. J. What is complex about complex disorders? Genome Biol. 13, 237 (2012).

8. Faraone, S. V. & Larsson, H. Genetics of attention deficit hyperactivity disorder. Mol. Psychiatry 24, 562–575 (2019).

9. Hilker, R. et al. Heritability of Schizophrenia and Schizophrenia Spectrum Based on the Nationwide Danish Twin Register. Biol. Psychiatry 83, 492–498 (2018).

10. Johansson, V., Kuja-Halkola, R., Cannon, T. D., Hultman, C. M. & Hedman, A. M. A population-based heritability estimate of bipolar disorder - In a Swedish twin sample. Psychiatry Res. 278, 180–187 (2019).

11. Kendler, K. S., Ohlsson, H., Lichtenstein, P., Sundquist, J. & Sundquist, K. The Genetic Epidemiology of Treated Major Depression in Sweden. Am. J. Psychiatry 175, 1137– 1144 (2018).

12. Andreassen, O. A., Hindley, G. F. L., Frei, O. & Smeland, O. B. New insights from the last decade of research in psychiatric genetics: discoveries, challenges and clinical implications. World Psychiatry Off. J. World Psychiatr. Assoc. WPA 22, 4–24 (2023).

13. van der Meer, D. et al. Mapping the genetic landscape of psychiatric disorders with the MiXeR toolset. Biol. Psychiatry (2025) doi:10.1016/j.biopsych.2025.02.886.

14. Hindley, G. et al. Charting the Landscape of Genetic Overlap Between Mental Disorders and Related Traits Beyond Genetic Correlation. Am. J. Psychiatry 179, 833–843 (2022).

15. Werme, J., van der Sluis, S., Posthuma, D. & de Leeuw, C. A. An integrated framework for local genetic correlation analysis. Nat. Genet. 54, 274–282 (2022).

16. Smeland, O. B. et al. The shared genetic risk architecture of neurological and psychiatric disorders: a genome-wide analysis. MedRxiv Prepr. Serv. Health Sci. 2023.07.21.23292993 (2023) doi:10.1101/2023.07.21.23292993.

17. Lee, P. H., Feng, Y.-C. A. & Smoller, J. W. Pleiotropy and Cross-Disorder Genetics Among Psychiatric Disorders. Biol. Psychiatry 89, 20–31 (2021).

18. Skou, S. T. et al. Multimorbidity. Nat. Rev. Dis. Primer 8, 48 (2022).

19. Halstead, S. et al. Prevalence of multimorbidity in people with and without severe mental illness: a systematic review and meta-analysis. Lancet Psychiatry 11, 431–442 (2024).

20. Momen, N. C. et al. Association between Mental Disorders and Subsequent Medical Conditions. N. Engl. J. Med. 382, 1721–1731 (2020).

21. Pizzol, D. et al. Relationship between severe mental illness and physical multimorbidity: a meta-analysis and call for action. *BMJ Ment*. Health 26, e300870 (2023).

22. De Hert, M. et al. Physical illness in patients with severe mental disorders. I. Prevalence, impact of medications and disparities in health care. World Psychiatry Off. J. World Psychiatr. Assoc. WPA 10, 52–77 (2011).

23. Plana-Ripoll, O. et al. A comprehensive analysis of mortality-related health metrics associated with mental disorders: a nationwide, register-based cohort study. The Lancet 394, 1827–1835 (2019).

24. Nordentoft, M. et al. Excess Mortality, Causes of Death and Life Expectancy in 270,770 Patients with Recent Onset of Mental Disorders in Denmark, Finland and Sweden. PLoS ONE 8, e55176 (2013).

25. Jayatilleke, N. et al. Contributions of specific causes of death to lost life expectancy in severe mental illness. Eur. Psychiatry J. Assoc. Eur. Psychiatr. 43, 109–115 (2017).

26. Li, L. et al. Attention-deficit/hyperactivity disorder as a risk factor for cardiovascular diseases: a nationwide population-based cohort study. World Psychiatry Off. J. World Psychiatr. Assoc. WPA 21, 452–459 (2022).

27. Du Rietz, E. et al. Mapping phenotypic and aetiological associations between ADHD and physical conditions in adulthood in Sweden: a genetically informed register study. Lancet Psychiatry 8, 774–783 (2021).

28. De Hert, M., Detraux, J. & Vancampfort, D. The intriguing relationship between coronary heart disease and mental disorders. Dialogues Clin. Neurosci. 20, 31–40 (2018).

29. Rødevand, L. et al. Characterizing the Shared Genetic Underpinnings of Schizophrenia and Cardiovascular Disease Risk Factors. Am. J. Psychiatry (2023) doi:10.1176/appi.ajp.20220660.

30. Bergstedt, J. et al. Distinct biological signature and modifiable risk factors underlie the comorbidity between major depressive disorder and cardiovascular disease. *Nat*. Cardiovasc. Res. 3, 754–769 (2024).

31. Bahrami, S. et al. Shared Genetic Loci Between Body Mass Index and Major Psychiatric Disorders: A Genome-wide Association Study. JAMA Psychiatry 77, 503–512 (2020).

32. Meijsen, J. et al. Quantifying the relative importance of genetics and environment on the comorbidity between mental and cardiometabolic disorders using 17 million Scandinavians. Nat. Commun. 15, 5064 (2024).

33. Hoogman, M. et al. Consortium neuroscience of attention deficit/hyperactivity disorder and autism spectrum disorder: The ENIGMA adventure. Hum. Brain Mapp. 43, 37–55 (2020).

34. van Erp, T. G. M. et al. Cortical Brain Abnormalities in 4474 Individuals With Schizophrenia and 5098 Control Subjects via the Enhancing Neuro Imaging Genetics Through Meta Analysis (ENIGMA) Consortium. Biol. Psychiatry 84, 644–654 (2018).

35. Hibar, D. P. et al. Cortical abnormalities in bipolar disorder: an MRI analysis of 6503 individuals from the ENIGMA Bipolar Disorder Working Group. Mol. Psychiatry 23, 932– 942 (2018).

36. Panizzon, M. S. et al. Distinct Genetic Influences on Cortical Surface Area and Cortical Thickness. Cereb. Cortex 19, 2728–2735 (2009).

37. Thompson, P. M. et al. ENIGMA and global neuroscience: A decade of large-scale studies of the brain in health and disease across more than 40 countries. Transl. Psychiatry 10, 1–28 (2020).

38. Sha, Z. et al. The overlapping genetic architecture of psychiatric disorders and cortical brain structure. BioRxiv Prepr. Serv. Biol. 2023.10.05.561040 (2023) doi:10.1101/2023.10.05.561040.

39. Cheng, W. et al. Shared genetic architecture between schizophrenia and subcortical brain volumes implicates early neurodevelopmental processes and brain development in childhood. Mol. Psychiatry 27, 5167–5176 (2022).

40. Grasby, K. L. et al. The genetic architecture of the human cerebral cortex. Science 367, eaay6690 (2020).

41. Gurholt, T. P. et al. Population-based body–brain mapping links brain morphology with anthropometrics and body composition. Transl. Psychiatry 11, 1–12 (2021).

42. Dove, A. et al. Cardiometabolic disease, cognitive decline, and brain structure in middle and older age. Alzheimers Dement. Diagn. Assess. Dis. Monit. 16, e12566 (2024).

43. Zhukovsky, P. et al. Genetic influences on brain and cognitive health and their interactions with cardiovascular conditions and depression. Nat. Commun. 15, 5207 (2024).

44. Antal, B. et al. Type 2 diabetes mellitus accelerates brain aging and cognitive decline: Complementary findings from UK Biobank and meta-analyses. eLife 11, e73138 (2022).

45. Di Cesare, M. et al. The Heart of the World. Glob. Heart 19, 11 (2024).

46. Shadrin, A. A. et al. Dissecting the genetic overlap between three complex phenotypes with trivariate MiXeR. MedRxiv Prepr. Serv. Health Sci. 2024.02.23.24303236 (2024) doi:10.1101/2024.02.23.24303236.

47. Sha, Z., Wager, T. D., Mechelli, A. & He, Y. Common Dysfunction of Large-Scale Neurocognitive Networks Across Psychiatric Disorders. Biol. Psychiatry 85, 379–388 (2019).

48. Roelfs, D. et al. Shared genetic architecture between mental health and the brain functional connectome in the UK Biobank. BMC Psychiatry 23, 461 (2023).

49. Spreng, R. N. & Turner, G. R. The Shifting Architecture of Cognition and Brain Function in Older Adulthood. Perspect. Psychol. Sci. J. Assoc. Psychol. Sci. 14, 523–542 (2019).

50. Boedhoe, P. S. W. et al. Subcortical Brain Volume, Regional Cortical Thickness, and Cortical Surface Area Across Disorders: Findings From the ENIGMA ADHD, ASD, and OCD Working Groups. Am. J. Psychiatry 177, 834–843 (2020).

51. Hoogman, M. et al. Brain Imaging of the Cortex in ADHD: A Coordinated Analysis of Large-Scale Clinical and Population-Based Samples. Am. J. Psychiatry 176, 531–542 (2019).

52. Sha, Z. et al. The overlapping genetic architecture of psychiatric disorders and cortical brain structure. BioRxiv Prepr. Serv. Biol. 2023.10.05.561040 (2023) doi:10.1101/2023.10.05.561040.

53. van der Meer, D. et al. Boosting Schizophrenia Genetics by Utilizing Genetic Overlap With Brain Morphology. Biol. Psychiatry 92, 291–298 (2022).

54. Lin, B. D. et al. Dissecting causal relationships between cortical morphology and neuropsychiatric disorders: a bidirectional Mendelian randomization study. Nat. Ment. Health 1–13 (2025) doi:10.1038/s44220-025-00397-4.

55. Chiappelli, J. et al. Tryptophan Metabolism and White Matter Integrity in Schizophrenia. Neuropsychopharmacology 41, 2587–2595 (2016).

56. van der Meer, D. et al. Atlas of plasma metabolic markers linked to human brain morphology. bioRxiv 2025.01.12.632645 (2025) doi:10.1101/2025.01.12.632645.

57. Beck, D. et al. Cardiometabolic risk factors associated with brain age and accelerate brain ageing. Hum. Brain Mapp. 43, 700–720 (2021).

58. Kang, H.-R., Kim, S. J., Nam, J. G., Park, Y. S. & Lee, C.-H. Impact of Smoking and Chronic Obstructive Pulmonary Disease on All-Cause, Respiratory, and Cardio-Cerebrovascular Mortality. Int. J. Chron. Obstruct. Pulmon. Dis. 19, 1261–1272 (2024).

59. Lv, H. et al. Association between Body Mass Index and Brain Health in Adults: A 16-Year Population-Based Cohort and Mendelian Randomization Study. Health Data Sci. 4, 0087 (2024).

60. Shang, X. et al. Association of a wide range of individual chronic diseases and their multimorbidity with brain volumes in the UK Biobank: A cross-sectional study. eClinicalMedicine 47, 101413 (2022).

61. Valletta, M. et al. Association of mild and complex multimorbidity with structural brain changes in older adults: A population-based study. Alzheimers Dement. 20, 1958–1965 (2024).

62. Ryan, C. M., van Duinkerken, E. & Rosano, C. Neurocognitive consequences of diabetes. Am. Psychol. 71, 563–576 (2016).

63. Locke, A. E. et al. Genetic studies of body mass index yield new insights for obesity biology. Nature 518, 197–206 (2015).

64. Finucane, H. K. et al. Partitioning heritability by functional annotation using genome-wide association summary statistics. Nat. Genet. 47, 1228–1235 (2015).

65. Morze, J. et al. Metabolomics and Type 2 Diabetes Risk: An Updated Systematic Review and Meta-analysis of Prospective Cohort Studies. Diabetes Care 45, 1013–1024 (2022).

66. Meer, D. van der et al. Divergent patterns of genetic overlap between severe mental disorders and metabolic markers. 2024.11.04.24316693 Preprint at 10.1101/2024.11.04.24316693 (2024).

67. Leppert, B. et al. The Effect of Attention Deficit/Hyperactivity Disorder on Physical Health Outcomes: A 2-Sample Mendelian Randomization Study. Am. J. Epidemiol. 190, 1047– 1055 (2021).

68. Thapar, A., Livingston, L. A., Eyre, O. & Riglin, L. Practitioner Review: Attention-deficit hyperactivity disorder and autism spectrum disorder - the importance of depression. J. Child Psychol. Psychiatry 64, 4–15 (2023).

69. Postolache, T. T. et al. Co-shared genetics and possible risk gene pathway partially explain the comorbidity of schizophrenia, major depressive disorder, type 2 diabetes, and metabolic syndrome. Am. J. Med. Genet. Part B Neuropsychiatr. Genet. Off. Publ. Int. Soc. Psychiatr. Genet. 180, 186–203 (2019).

70. Ding, H. et al. Shared genetics of psychiatric disorders and type 2 diabetes:a large-scale genome-wide cross-trait analysis. J. Psychiatr. Res. 159, 185–195 (2023).

71. Predescu, E., Vaidean, T., Rapciuc, A.-M. & Sipos, R. Metabolomic Markers in Attention-Deficit/Hyperactivity Disorder (ADHD) among Children and Adolescents—A Systematic Review. Int. J. Mol. Sci. 25, 4385 (2024).

72. Dehnavi, A. Z. et al. Association of ADHD symptoms with type 2 diabetes and cardiovascular comorbidities in adults receiving outpatient diabetes care. J. Clin. Transl. Endocrinol. 32, 100318 (2023).

73. Trubetskoy, V. et al. Mapping genomic loci implicates genes and synaptic biology in schizophrenia. Nature 604, 502–508 (2022).

74. Demontis, D. et al. Genome-wide analyses of ADHD identify 27 risk loci, refine the genetic architecture and implicate several cognitive domains. Nat. Genet. 55, 198–208 (2023).

75. Suzuki, K. et al. Genetic drivers of heterogeneity in type 2 diabetes pathophysiology. Nature 627, 347–357 (2024).

76. Aragam, K. G. et al. Discovery and systematic characterization of risk variants and genes for coronary artery disease in over a million participants. Nat. Genet. 54, 1803– 1815 (2022).

77. Sudlow, C. et al. UK Biobank: An Open Access Resource for Identifying the Causes of a Wide Range of Complex Diseases of Middle and Old Age. PLoS Med. 12, e1001779 (2015).

78. Manichaikul, A. et al. Robust relationship inference in genome-wide association studies. Bioinforma. Oxf. Engl. 26, 2867–2873 (2010).

79. Bulik-Sullivan, B. K. et al. LD Score regression distinguishes confounding from polygenicity in genome-wide association studies. Nat. Genet. 47, 291–295 (2015).

80. Benjamini, Y. & Yekutieli, D. The Control of the False Discovery Rate in Multiple Testing under Dependency. Ann. Stat. 29, 1165–1188 (2001).

81. Smeland, O. B. et al. Discovery of shared genomic loci using the conditional false discovery rate approach. Hum. Genet. 139, 85–94 (2020).

82. Holland, D. et al. Beyond SNP heritability: Polygenicity and discoverability of phenotypes estimated with a univariate Gaussian mixture model. PLoS Genet. 16, e1008612 (2020).

83. Frei, O. et al. Bivariate causal mixture model quantifies polygenic overlap between complex traits beyond genetic correlation. Nat. Commun. 10, 2417 (2019).

84. Bowden, J., Davey Smith, G., Haycock, P. C. & Burgess, S. Consistent Estimation in Mendelian Randomization with Some Invalid Instruments Using a Weighted Median Estimator. Genet. Epidemiol. 40, 304–314 (2016).

85. Burgess, S. & Thompson, S. G. Interpreting findings from Mendelian randomization using the MR-Egger method. Eur. J. Epidemiol. 32, 377–389 (2017).

86. Hartwig, F. P., Davey Smith, G. & Bowden, J. Robust inference in summary data Mendelian randomization via the zero modal pleiotropy assumption. Int. J. Epidemiol. 46, 1985–1998 (2017).

87. Andreassen, O. A. et al. Improved Detection of Common Variants Associated with Schizophrenia and Bipolar Disorder Using Pleiotropy-Informed Conditional False Discovery Rate. PLOS Genet. 9, e1003455 (2013).

88. Rødevand, L. et al. Polygenic overlap and shared genetic loci between loneliness, severe mental disorders, and cardiovascular disease risk factors suggest shared molecular mechanisms. Transl. Psychiatry 11, 3 (2021).

89. Mountjoy, E. et al. An open approach to systematically prioritize causal variants and genes at all published human GWAS trait-associated loci. Nat. Genet. 53, 1527–1533 (2021).

90. Sherman, B. T. et al. DAVID: a web server for functional enrichment analysis and functional annotation of gene lists (2021 update). Nucleic Acids Res. 50, W216–W221 (2022).

91. Huang, D. W., Sherman, B. T. & Lempicki, R. A. Systematic and integrative analysis of large gene lists using DAVID bioinformatics resources. Nat. Protoc. 4, 44–57 (2009).

92. Kanehisa, M. & Goto, S. KEGG: Kyoto Encyclopedia of Genes and Genomes. Nucleic Acids Res. 28, 27–30 (2000).

93. Milacic, M. et al. The Reactome Pathway Knowledgebase 2024. Nucleic Acids Res. 52, D672–D678 (2024).

94. Grove, J. et al. Identification of common genetic risk variants for autism spectrum disorder. Nat. Genet. 51, 431–444 (2019).

95. O’Connell, K. S. et al. Genomics yields biological and phenotypic insights into bipolar disorder. Nature 639, 968–975 (2025).

96. Adams, M. J. et al. Trans-ancestry genome-wide study of depression identifies 697 associations implicating cell types and pharmacotherapies. Cell 188, 640–652.e9 (2025).

