## Supplementary Materials for "Mapping genetic convergence across brain structure, mental health, and cardiometabolic disease"

### Supplementary Material

#### Supplementary Figures

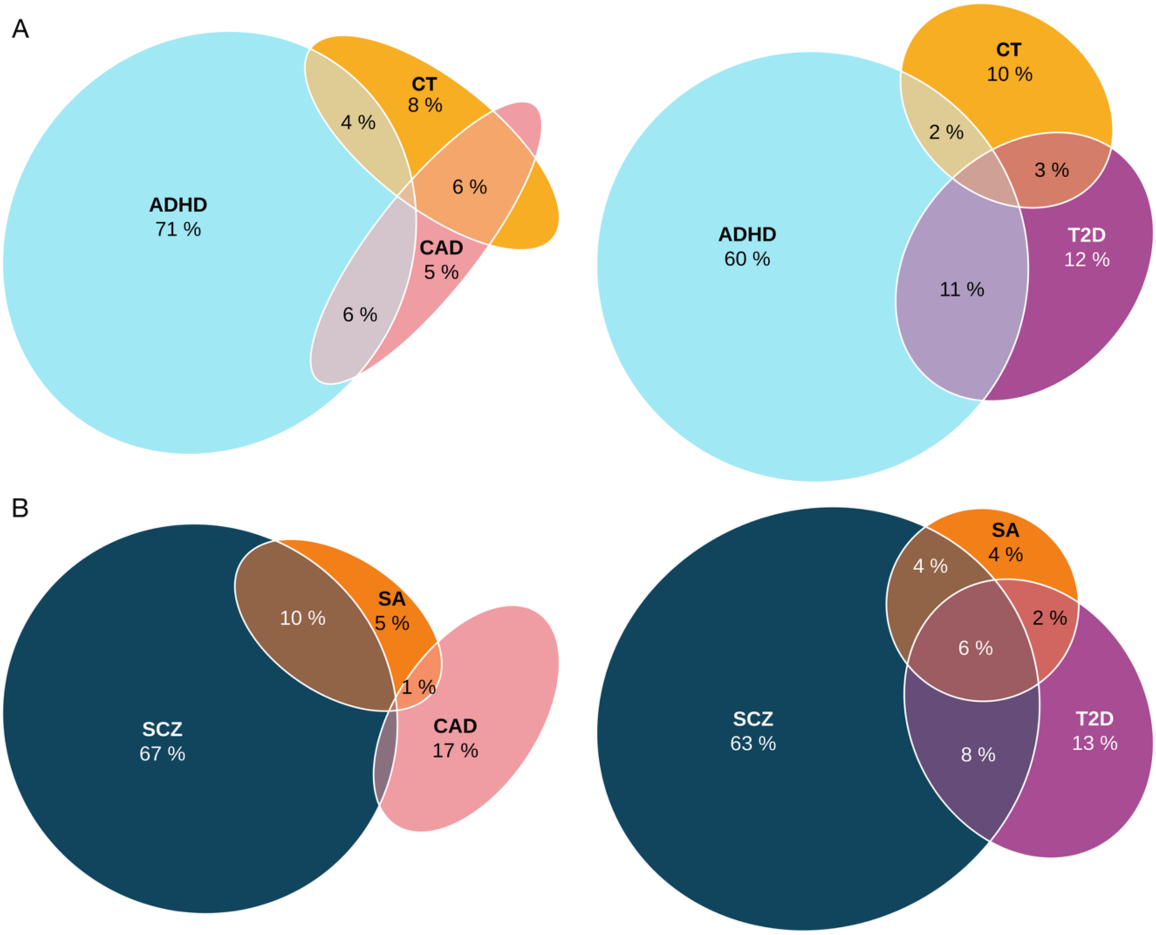


**Supplementary Figure 1: Complementary trivariate overlap between psychiatric disorders, cardiometabolic disease and cortical morphology**
**A.** Genetic overlap between ADHD, CT, and cardiometabolic disease. Based on trivariate MiXeR analyses, the Venn diagrams illustrate the extent of genetic overlap between cortical morphology (i.e., CT), ADHD, and cardiometabolic traits (CAD and T2D). Only genetic overlaps exceeding 1% are annotated. **B.** Genetic overlap between SCZ, SA, and cardiometabolic disease. Genetic overlaps between CT, SCZ, and the cardiometabolic traits CAD and T2D are presented using Venn diagrams.

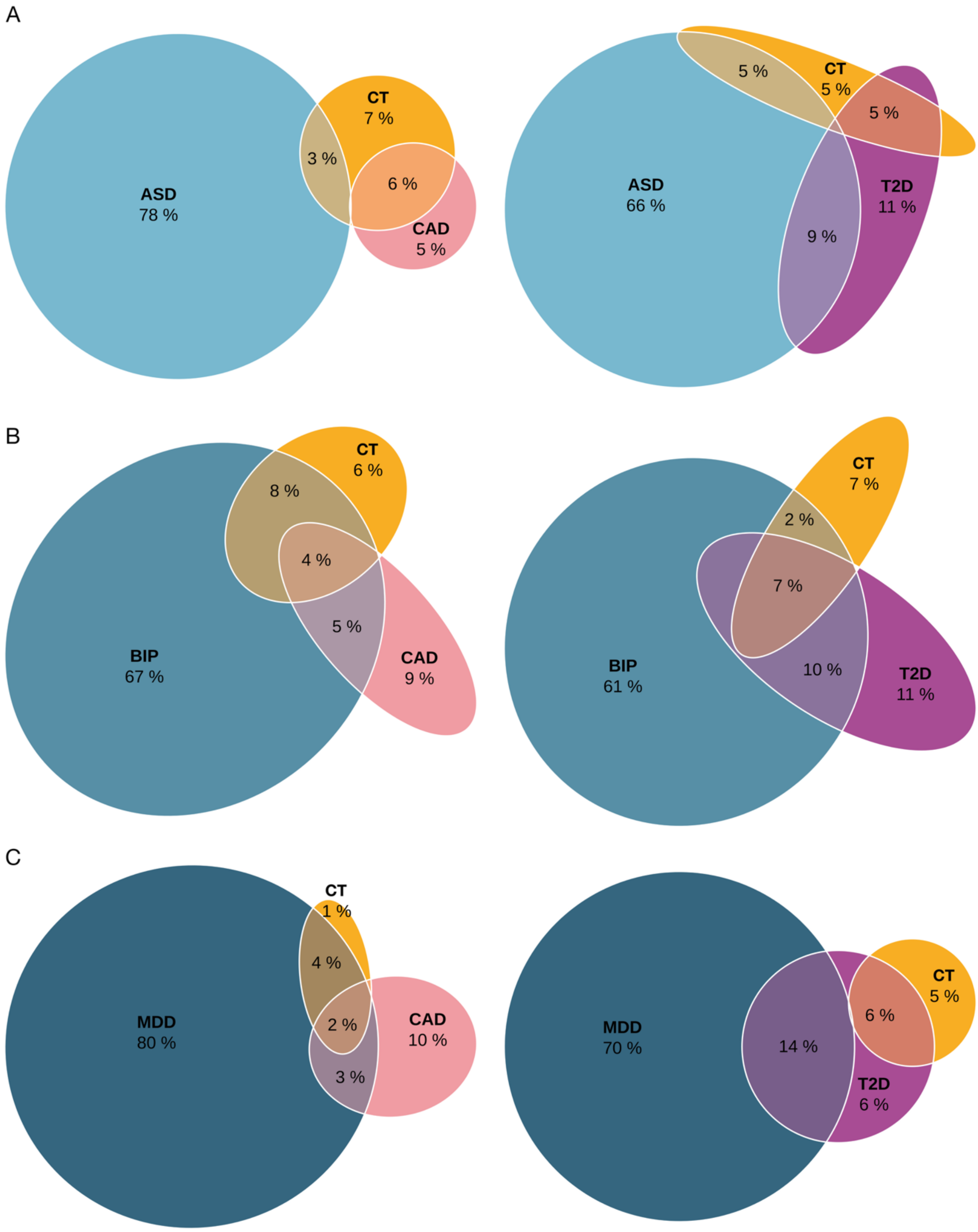


**Supplementary Figure 2: Trivariate overlap between remaining psychiatric disorders, cardiometabolic disease and cortical thickness**

Genetic overlap between three psychiatric disorders, CT, and cardiometabolic disease. Based on trivariate MiXeR analyses, the Venn diagrams illustrate the extent of genetic overlap between cortical morphology (i.e., CT), psychiatric disorder (i.e., ASD, BIP, MDD), and cardiometabolic traits (CAD and T2D). Only genetic overlaps exceeding 1% are annotated. Results are presented for ASD (**A**), BIP (**B**), and MDD (**C**).

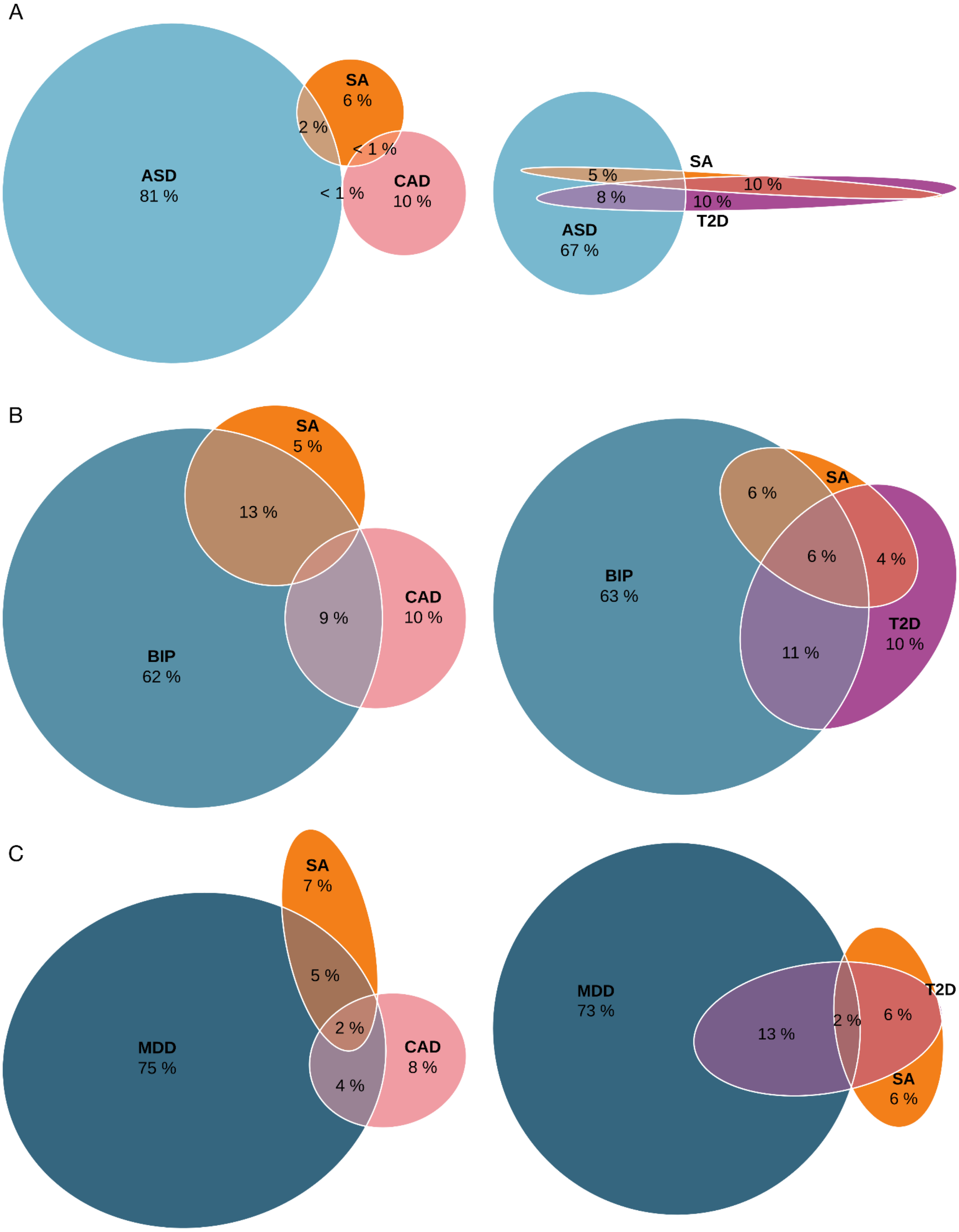


**Supplementary Figure 3: Trivariate overlap between remaining psychiatric disorders, cardiometabolic disease and surface area**

Genetic overlap between three psychiatric disorders, SA, and cardiometabolic disease. Based on trivariate MiXeR analyses, the Venn diagrams illustrate the extent of genetic overlap between cortical morphology (i.e., SA), psychiatric disorder (i.e., ASD, BIP, MDD), and cardiometabolic traits (CAD and T2D). Only genetic overlaps exceeding 1% are annotated. Results are presented for ASD (**A**), BIP (**B**), and MDD (**C**).

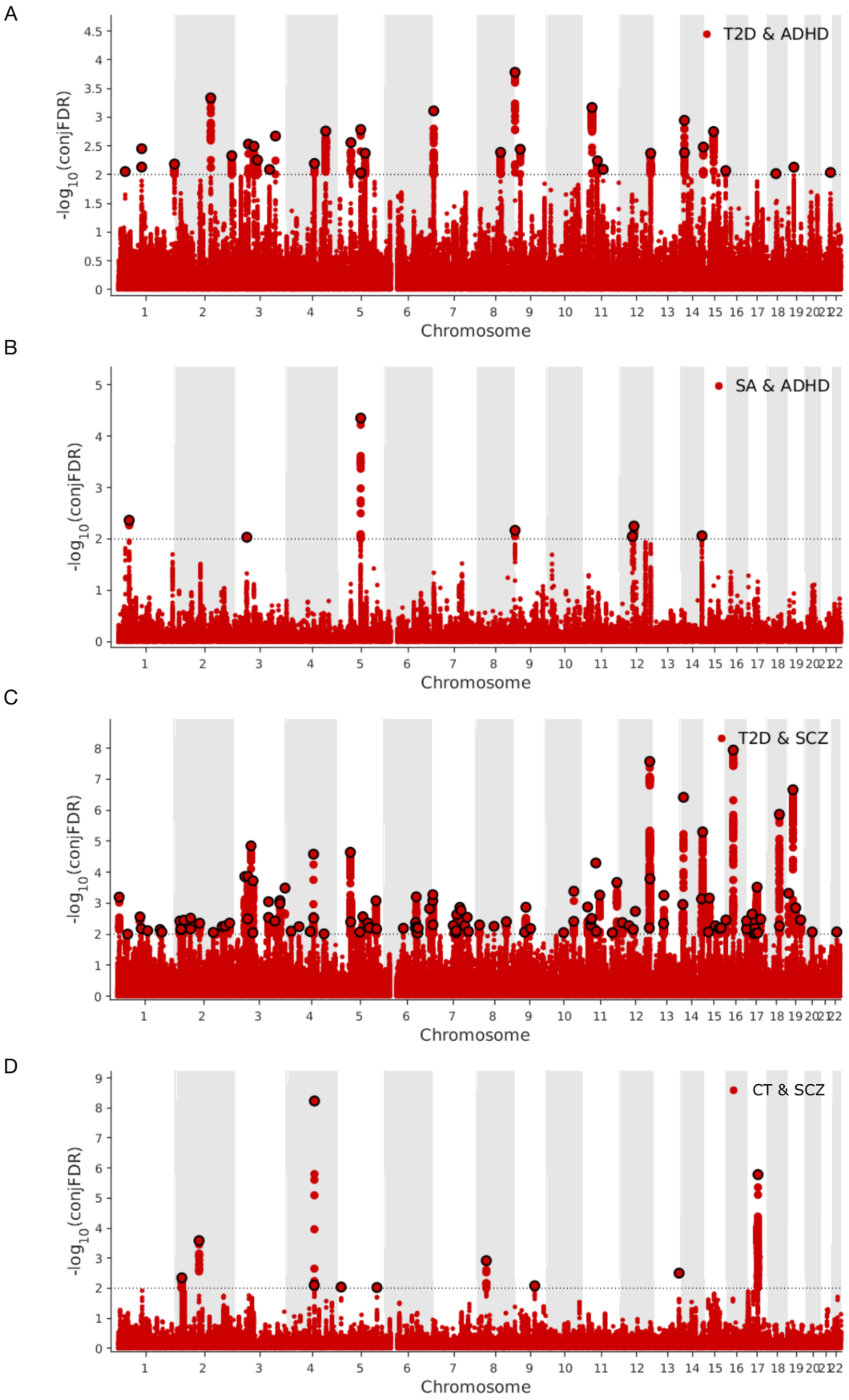


**Supplementary Figure 4: Shared loci at conjDFR < 0.01**

We searched for SNPs jointly associated between ADHD and SA (**A**), ADHD and T2D (**B**), SCZ and T2D (**C**), as well as SCZ and CT (**D**). For these four analyses, the associated Manhattan plot displays genetic variants shared by the respective two traits. Each Manhattan plot shows the $-\log_{10}$ transformed conjFDR values for each SNP on the y-axis. SNPs with conjFDR < 0.01are shown with enlarged data points. A black circle around the enlarged data points indicates the most significant SNP in each linkage disequilibrium block. The analysis revealed 30, 7, 110, and 9 lead SNPs respectively.

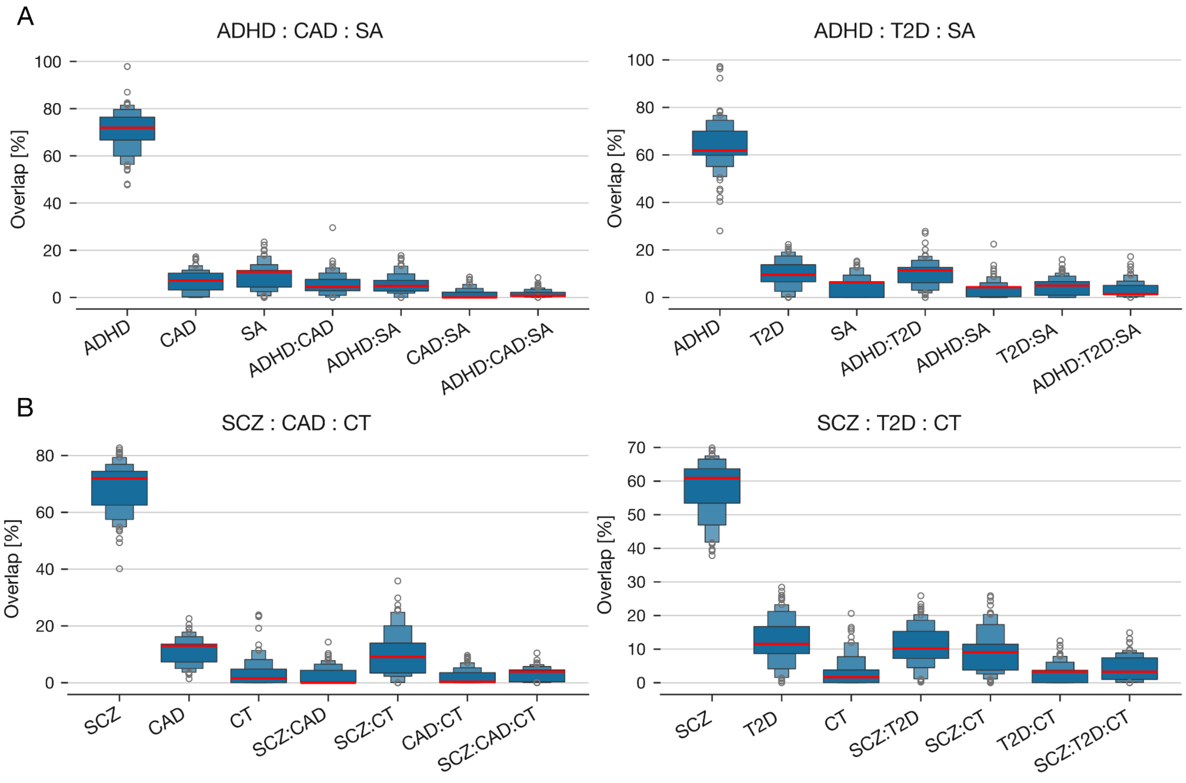


**Supplementary Figure 5: Stability of trivariate MiXeR model estimates across optimization runs**

The trivariate MiXeR results presented in the main text reflect optimal estimates derived from 100 independent optimization runs. For each polygenicity parameter, the median value across runs was computed, and the final result was selected as the run with the smallest deviation from the median overlap pattern. Each panel displays boxen plots of polygenicity and pairwise overlap estimates derived from 100 independent model fits (**A** for ADHD, **B** for SCZ). Boxen plots depict multiple nested quantile ranges (e.g., deciles), enabling a high-resolution view of the distribution tails and skewness beyond what standard box plots capture. The optimal run (used in the main text) is highlighted in red.

#### Supplementary Tables

**Supplementary Table 1:** Mendelian Randomization results
The table summarizes Mendelian Randomization (MR) estimates for each exposure–outcome pair tested using different methods implemented in the TwoSampleMR framework. Columns report the exposure and outcome traits, the MR method used, the number of SNPs included as instruments (nsnp), the estimated causal effect (b), its standard error (se), and the corresponding p-value (pval). Results reflect harmonized and clumped datasets based on genome-wide summary statistics.

| Outcome | Exposure | Method | nsnp | b | se | pval |
|---|---|---|---|---|---|---|
| T2D | ADHD | MR Egger | 25 | -0,130 | 0,158 | 0,419 |
| T2D | ADHD | Weighted median | 25 | 0,076 | 0,024 | 0,002 |
| T2D | ADHD | Inverse variance weighted | 25 | 0,072 | 0,035 | 0,041 |
| T2D | ADHD | Simple mode | 25 | 0,027 | 0,064 | 0,675 |
| T2D | ADHD | Weighted mode | 25 | 0,048 | 0,039 | 0,228 |
| T2D | SCZ | MR Egger | 148 | -0,005 | 0,065 | 0,942 |
| T2D | SCZ | Weighted median | 148 | -0,020 | 0,010 | 0,047 |
| T2D | SCZ | Inverse variance weighted | 148 | -0,012 | 0,014 | 0,400 |
| T2D | SCZ | Simple mode | 148 | -0,036 | 0,028 | 0,202 |
| T2D | SCZ | Weighted mode | 148 | -0,033 | 0,022 | 0,137 |
| T2D | CT | MR Egger | 21 | -0,123 | 0,174 | 0,488 |
| T2D | CT | Weighted median | 21 | -0,004 | 0,036 | 0,907 |
| T2D | CT | Inverse variance weighted | 21 | -0,091 | 0,054 | 0,090 |
| T2D | CT | Simple mode | 21 | 0,032 | 0,066 | 0,632 |
| T2D | CT | Weighted mode | 21 | 0,029 | 0,045 | 0,521 |
| ADHD | T2D | MR Egger | 423 | 0,010 | 0,041 | 0,817 |
| ADHD | T2D | Weighted median | 423 | 0,068 | 0,022 | 0,002 |
| ADHD | T2D | Inverse variance weighted | 423 | 0,128 | 0,019 | 1,4E-11 |
| ADHD | T2D | Simple mode | 423 | 0,089 | 0,067 | 0,184 |

| | | | | | | |
|---|---|---|---|---|---|---|
| ADHD | T2D | Weighted mode | 423 | 0,066 | 0,037 | 0,078 |
| SCZ | T2D | MR Egger | 454 | 0,002 | 0,044 | 0,957 |
| SCZ | T2D | Weighted median | 454 | -0,005 | 0,023 | 0,826 |
| SCZ | T2D | Inverse variance weighted | 454 | -0,033 | 0,020 | 0,108 |
| SCZ | T2D | Simple mode | 454 | -0,059 | 0,062 | 0,343 |
| SCZ | T2D | Weighted mode | 454 | -0,004 | 0,033 | 0,908 |
| SCZ | CT | MR Egger | 21 | 0,233 | 0,451 | 0,611 |
| SCZ | CT | Weighted median | 21 | 0,111 | 0,080 | 0,166 |
| SCZ | CT | Inverse variance weighted | 21 | 0,113 | 0,138 | 0,412 |
| SCZ | CT | Simple mode | 21 | 0,034 | 0,146 | 0,821 |
| SCZ | CT | Weighted mode | 21 | 0,050 | 0,118 | 0,676 |
| SA | T2D | MR Egger | 455 | 0,038 | 0,025 | 0,121 |
| SA | T2D | Weighted median | 455 | -0,024 | 0,014 | 0,097 |
| SA | T2D | Inverse variance weighted | 455 | -0,018 | 0,012 | 0,117 |
| SA | T2D | Simple mode | 455 | -0,011 | 0,035 | 0,755 |
| SA | T2D | Weighted mode | 455 | -0,021 | 0,019 | 0,265 |
| SA | ADHD | MR Egger | 25 | 0,167 | 0,128 | 0,204 |
| SA | ADHD | Weighted median | 25 | 0,021 | 0,031 | 0,492 |
| SA | ADHD | Inverse variance weighted | 25 | -0,031 | 0,029 | 0,289 |
| SA | ADHD | Simple mode | 25 | 0,031 | 0,058 | 0,594 |
| SA | ADHD | Weighted mode | 25 | 0,036 | 0,057 | 0,537 |
| T2D | SA | MR Egger | 22 | 0,156 | 0,288 | 0,595 |
| T2D | SA | Weighted median | 22 | -0,085 | 0,043 | 0,049 |
| T2D | SA | Inverse variance weighted | 22 | -0,015 | 0,104 | 0,883 |
| T2D | SA | Simple mode | 22 | -0,078 | 0,059 | 0,203 |
| T2D | SA | Weighted mode | 22 | -0,132 | 0,033 | 0,001 |

| | | | | | | |
|---|---|---|---|---|---|---|
| ADHD | SA | MR Egger | 20 | -0,205 | 0,182 | 0,275 |
| ADHD | SA | Weighted median | 20 | -0,231 | 0,071 | 0,001 |
| ADHD | SA | Inverse variance weighted | 20 | -0,260 | 0,067 | 9,6E-05 |
| ADHD | SA | Simple mode | 20 | -0,215 | 0,113 | 0,072 |
| ADHD | SA | Weighted mode | 20 | -0,239 | 0,079 | 0,007 |
| CT | T2D | MR Egger | 455 | -0,011 | 0,023 | 0,622 |
| CT | T2D | Weighted median | 455 | -0,022 | 0,016 | 0,152 |
| CT | T2D | Inverse variance weighted | 455 | -0,030 | 0,011 | 0,005 |
| CT | SCZ | MR Egger | 148 | -0,143 | 0,066 | 0,032 |
| CT | SCZ | Weighted median | 148 | -0,035 | 0,015 | 0,022 |
| CT | SCZ | Inverse variance weighted | 148 | -0,010 | 0,015 | 0,493 |

**Supplementary Table 2: Genes identified across conjFDR analyses**
This table lists all genes mapped from significant SNPs identified in the conjunctional FDR (conjFDR) analyses across the tested trait pairs. For each gene, the corresponding Ensembl gene ID and official gene symbol are provided.

| ADHD & T2D | | ADHD & SA | | SCZ & T2D | | SCZ & CT | |
|---|---|---|---|---|---|---|---|
| Ensembl ID | Symbol | Ensembl ID | Symbol | Ensembl ID | Symbol | Ensembl ID | Symbol |
| ENSG00000204120 | *GIGYF2* | ENSG00000126091 | *ST3GAL3* | ENSG00000041988 | *THAP3* | ENSG00000213699 | *SLC35F6* |
| ENSG00000115474 | *KCNJ13* | ENSG00000117020 | *AKT3* | ENSG00000168653 | *NDUFS5* | ENSG00000138030 | *KHK* |
| ENSG00000182600 | *SNORC* | ENSG00000168036 | *CTNNB1* | ENSG00000090621 | *PABPC4* | ENSG00000157851 | *DPYSL5* |
| ENSG00000093167 | *LRRFIP2* | ENSG00000160972 | *PPP1R16A* | ENSG00000204060 | *FOXO6* | ENSG00000138028 | *CGREF1* |
| ENSG00000164068 | *RNF123* | ENSG00000160957 | *RECQL4* | ENSG00000162433 | *AK4* | ENSG00000205221 | *VIT* |
| ENSG00000004534 | *RBM6* | ENSG00000134285 | *FKBP11* | ENSG00000153904 | *DDAH1* | ENSG00000158158 | *CNNM4* |
| ENSG00000164078 | *MST1R* | ENSG00000181929 | *PRKAG1* | ENSG00000007341 | *ST7L* | ENSG00000115073 | *ACTR1B* |
| ENSG00000001617 | *SEMA3F* | ENSG00000181418 | *DDN* | ENSG00000122218 | *COPA* | ENSG00000196141 | *SPATS2L* |
| ENSG00000158234 | *FAIM* | ENSG00000139636 | *LMBR1L* | ENSG00000162736 | *NCSTN* | ENSG00000168268 | *NT5DC2* |
| ENSG00000158186 | *MRAS* | ENSG00000111540 | *RAB5B* | ENSG00000184144 | *CNTN2* | ENSG00000055955 | *ITIH4* |
| ENSG00000109323 | *MANBA* | ENSG00000139531 | *SUOX* | ENSG00000117335 | *CD46* | ENSG00000163939 | *PBRM1* |
| ENSG00000164037 | *SLC9B1* | ENSG00000120860 | *WASHC3* | ENSG00000138030 | *KHK* | ENSG00000016864 | *GLT8D1* |
| ENSG00000109332 | *UBE2D3* | ENSG00000150967 | *ABCB9* | ENSG00000157851 | *DPYSL5* | ENSG00000055957 | *ITIH1* |
| ENSG00000145354 | *CISD2* | ENSG00000051825 | *MPHOSPH9* | ENSG00000138028 | *CGREF1* | ENSG00000162267 | *ITIH3* |
| ENSG00000164167 | *LSM6* | ENSG00000183955 | *KMT5A* | ENSG00000157992 | *KRTCAP3* | ENSG00000109572 | *CLCN3* |
| ENSG00000172262 | *ZNF131* | | | ENSG00000138002 | *IFT172* | ENSG00000055147 | *FAM114A2* |
| ENSG00000002822 | *MAD1L1* | | | ENSG00000115226 | *FNDC4* | ENSG00000171097 | *KYAT1* |
| ENSG00000160972 | *PPP1R16A* | | | ENSG00000243147 | *MRPL33* | ENSG00000119689 | *DLST* |
| ENSG00000160957 | *RECQL4* | | | ENSG00000162869 | *PPP1R21* | ENSG00000198208 | *RPS6KL1* |
| ENSG00000164989 | *CCDC171* | | | ENSG00000136536 | *MARCHF7* | ENSG00000140474 | *ULK3* |
| ENSG00000187164 | *SHTN1* | | | ENSG00000241399 | *CD302* | ENSG00000140506 | *LMAN1L* |

| | |
|---|---|
| ENSG00000070081 | *NUCB2* |
| ENSG00000187486 | *KCNJ11* |
| ENSG00000080854 | *IGSF9B* |
| ENSG00000079387 | *SENP1* |
| ENSG00000152556 | *PFKM* |
| ENSG00000167528 | *ZNF641* |
| ENSG00000181929 | *PRKAG1* |
| ENSG00000120860 | *WASHC3* |
| ENSG00000150967 | *ABCB9* |
| ENSG00000051825 | *MPHOSPH9* |
| ENSG00000183955 | *KMT5A* |
| ENSG00000103227 | *LMF1* |
| ENSG00000188554 | *NBR1* |
| ENSG00000012048 | *BRCA1* |
| ENSG00000133275 | *CSNK1G2* |
| ENSG00000064490 | *RFXANK* |
| ENSG00000254901 | *BORCS8* |
| ENSG00000129933 | *MAU2* |
| ENSG00000167491 | *GATAD2A* |
| ENSG00000197381 | *ADARB1* |
| ENSG00000186866 | *POFUT2* |

| | |
|---|---|
| ENSG00000154479 | *CFAP210* |
| ENSG00000138430 | *OLA1* |
| ENSG00000152430 | *BOLL* |
| ENSG00000115896 | *PLCL1* |
| ENSG00000153827 | *TRIP12* |
| ENSG00000134077 | *THUMPD3* |
| ENSG00000093167 | *LRRFIP2* |
| ENSG00000164077 | *MON1A* |
| ENSG00000164068 | *RNF123* |
| ENSG00000004534 | *RBM6* |
| ENSG00000164078 | *MST1R* |
| ENSG00000173531 | *MST1* |
| ENSG00000168268 | *NT5DC2* |
| ENSG00000055955 | *ITIH4* |
| ENSG00000213533 | *STIMATE* |
| ENSG00000055957 | *ITIH1* |
| ENSG00000162267 | *ITIH3* |
| ENSG00000272573 | *MUSTN1* |
| ENSG00000163933 | *RFT1* |
| ENSG00000114054 | *PCCB* |
| ENSG00000168917 | *SLC35G2* |
| ENSG00000156931 | *VPS8* |
| ENSG00000090316 | *MAEA* |
| ENSG00000159692 | *CTBP1* |
| ENSG00000163945 | *UVSSA* |
| ENSG00000132405 | *TBC1D14* |

| | |
|---|---|
| ENSG00000178802 | *MPI* |
| ENSG00000108953 | *YWHAE* |
| ENSG00000132388 | *UBE2G1* |
| ENSG00000186868 | *MAPT* |
| ENSG00000120071 | *KANSL1* |
| ENSG00000225190 | *PLEKHM1* |
| ENSG00000120088 | *CRHR1* |
| ENSG00000159314 | *ARHGAP27* |
| ENSG00000185294 | *SPPL2C* |
| ENSG00000238083 | *LRRC37A2* |
| ENSG00000185829 | *ARL17A* |

| | |
|---|---|
| ENSG00000145335 | *SNCA* |
| ENSG00000109323 | *MANBA* |
| ENSG00000109332 | *UBE2D3* |
| ENSG00000145354 | *CISD2* |
| ENSG00000164037 | *SLC9B1* |
| ENSG00000164305 | *CASP3* |
| ENSG00000151725 | *CENPU* |
| ENSG00000145730 | *PAM* |
| ENSG00000205359 | *SLCO6A1* |
| ENSG00000145779 | *TNFAIP8* |
| ENSG00000133835 | *HSD17B4* |
| ENSG00000213523 | *SRA1* |
| ENSG00000113108 | *APBB3* |
| ENSG00000170458 | *CD14* |
| ENSG00000113119 | *TMCO6* |
| ENSG00000055147 | *FAM114A2* |
| ENSG00000037749 | *MFAP3* |
| ENSG00000146285 | *SCML4* |
| ENSG00000047932 | *GOPC* |
| ENSG00000111879 | *FAM184A* |
| ENSG00000002822 | *MAD1L1* |
| ENSG00000122687 | *MRM2* |
| ENSG00000188732 | *FAM221A* |
| ENSG00000087077 | *TRIP6* |
| ENSG00000135250 | *SRPK2* |
| ENSG00000105894 | *PTN* |

| | |
|---|---|
| ENSG00000175806 | *MSRA* |
| ENSG00000154359 | *LONRF1* |
| ENSG00000104447 | *TRPS1* |
| ENSG00000171045 | *TSNARE1* |
| ENSG00000176956 | *LY6H* |
| ENSG00000198876 | *DCAF12* |
| ENSG00000095383 | *TBC1D2* |
| ENSG00000070061 | *ELP1* |
| ENSG00000136877 | *FPGS* |
| ENSG00000166272 | *WBP1L* |
| ENSG00000138111 | *MFSD13A* |
| ENSG00000173915 | *ATP5MK* |
| ENSG00000070081 | *NUCB2* |
| ENSG00000187486 | *KCNJ11* |
| ENSG00000152219 | *ARL14EP* |
| ENSG00000109919 | *MTCH2* |
| ENSG00000172922 | *RNASEH2C* |
| ENSG00000172977 | *KAT5* |
| ENSG00000172757 | *CFL1* |
| ENSG00000172803 | *SNX32* |
| ENSG00000175334 | *BANF1* |
| ENSG00000172732 | *MUS81* |
| ENSG00000172500 | *FIBP* |
| ENSG00000162341 | *TPCN2* |
| ENSG00000149292 | *TTC12* |
| ENSG00000120451 | *SNX19* |

| | |
|---|---|
| ENSG00000080854 | *IGSF9B* |
| ENSG00000135409 | *AMHR2* |
| ENSG00000123329 | *ARHGAP9* |
| ENSG00000179912 | *R3HDM2* |
| ENSG00000089041 | *P2RX7* |
| ENSG00000111011 | *RSRC2* |
| ENSG00000051825 | *MPHOSPH9* |
| ENSG00000150967 | *ABCB9* |
| ENSG00000183955 | *KMT5A* |
| ENSG00000119242 | *CCDC92* |
| ENSG00000197653 | *DNAH10* |
| ENSG00000179195 | *ZNF664* |
| ENSG00000196498 | *NCOR2* |
| ENSG00000122034 | *GTF3A* |
| ENSG00000151773 | *CCDC122* |
| ENSG00000151327 | *FAM177A1* |
| ENSG00000100883 | *SRP54* |
| ENSG00000126777 | *KTN1* |
| ENSG00000100731 | *PCNX1* |
| ENSG00000075413 | *MARK3* |
| ENSG00000126214 | *KLC1* |
| ENSG00000166165 | *CKB* |
| ENSG00000256053 | *COA8* |
| ENSG00000166166 | *TRMT61A* |
| ENSG00000156414 | *TDRD9* |
| ENSG00000088808 | *PPP1R13B* |

| | |
|---|---|
| ENSG00000100711 | *ZFYVE21* |
| ENSG00000168803 | *MAPDA* |
| ENSG00000104055 | *TGM5* |
| ENSG00000159495 | *TGM7* |
| ENSG00000140265 | *ZSCAN29* |
| ENSG00000166762 | *CATSPER2* |
| ENSG00000242866 | *STRC* |
| ENSG00000136378 | *ADAMTS7* |
| ENSG00000103876 | *FAH* |
| ENSG00000136383 | *ALPK3* |
| ENSG00000182511 | *FES* |
| ENSG00000153406 | *NMRAL1* |
| ENSG00000089486 | *CDIP1* |
| ENSG00000103423 | *DNAJA3* |
| ENSG00000103415 | *HMOX2* |
| ENSG00000169592 | *INO80E* |
| ENSG00000149927 | *DOC2A* |
| ENSG00000149926 | *TLCD3B* |
| ENSG00000090238 | *YPEL3* |
| ENSG00000102886 | *GDPD3* |
| ENSG00000005844 | *ITGAL* |
| ENSG00000159593 | *NAE1* |
| ENSG00000141076 | *UTP4* |
| ENSG00000040199 | *PHLPP2* |
| ENSG00000153774 | *CFDP1* |
| ENSG00000197912 | *SPG7* |

| | |
|---|---|
| ENSG00000178773 | *CPNE7* |
| ENSG00000158792 | *SPATA2L* |
| ENSG00000185324 | *CDK10* |
| ENSG00000187741 | *FANCA* |
| ENSG00000167523 | *SPATA33* |
| ENSG00000131165 | *CHMP1A* |
| ENSG00000158805 | *ZNF276* |
| ENSG00000075399 | *VPS9D1* |
| ENSG00000074755 | *ZZEF1* |
| ENSG00000132388 | *UBE2G1* |
| ENSG00000161929 | *SCIMP* |
| ENSG00000108559 | *NUP88* |
| ENSG00000072778 | *ACADVL* |
| ENSG00000170291 | *ELP5* |
| ENSG00000175662 | *TOM1L2* |
| ENSG00000108557 | *RAI1* |
| ENSG00000072310 | *SREBF1* |
| ENSG00000171962 | *DRC3* |
| ENSG00000108671 | *PSMD11* |
| ENSG00000278259 | *MYO19* |
| ENSG00000278311 | *GGNBP2* |
| ENSG00000161395 | *PGAP3* |
| ENSG00000073605 | *GSDMB* |
| ENSG00000131748 | *STARD3* |
| ENSG00000186868 | *MAPT* |
| ENSG00000120071 | *KANSL1* |

| | |
|---|---|
| ENSG00000225190 | *PLEKHM1* |
| ENSG00000120088 | *CRHR1* |
| ENSG00000159314 | *ARHGAP27* |
| ENSG00000185294 | *SPPL2C* |
| ENSG00000238083 | *LRRC37A2* |
| ENSG00000185829 | *ARL17A* |
| ENSG00000159202 | *UBE2Z* |
| ENSG00000136436 | *CALCOCO2* |
| ENSG00000170703 | *TTLL6* |
| ENSG00000159210 | *SNF8* |
| ENSG00000159199 | *ATP5MC1* |
| ENSG00000159640 | *ACE* |
| ENSG00000055483 | *USP36* |
| ENSG00000122490 | *SLC66A2* |
| ENSG00000127663 | *KDM4B* |
| ENSG00000129933 | *MAU2* |
| ENSG00000167491 | *GATAD2A* |
| ENSG00000064490 | *RFXANK* |
| ENSG00000254901 | *BORCS8* |
| ENSG00000089639 | *GMIP* |
| ENSG00000064547 | *LPAR2* |
| ENSG00000105185 | *PDCD5* |
| ENSG00000176920 | *FUT2* |
| ENSG00000176909 | *MAMSTR* |
| ENSG00000078747 | *ITCH* |
| ENSG00000101460 | *MAP1LC3A* |

| | |
|---|---|
| ENSG00000131069 | *ACSS2* |
| ENSG00000100991 | *TRPC4AP* |
| ENSG00000088298 | *EDEM2* |
| ENSG00000100027 | *YPEL1* |
| ENSG00000159873 | *CCDC117* |
| ENSG00000172346 | *CSDC2* |
| ENSG00000100266 | *PACSIN2* |
| ENSG00000242247 | *ARFGAP3* |
| ENSG00000184164 | *CRELD2* |
| ENSG00000198355 | *PIM3* |

**Supplementary Table 3: Enriched biological pathways**

The table presents enriched pathways among genes of interest, with KEGG pathways highlighted in red and REACTOME pathways in blue. Reported columns include the number of input genes associated with each pathway (Count), the raw p-value from the modified Fisher's exact test, and the fold enrichment. Fold enrichment reflects the ratio between the proportion of genes associated with a given pathway in the input list and the corresponding proportion in the background population. Representative genes are shown for each term.

**ADHD & T2D**

| Term | Count | p-value | Genes | Fold Enrichment |
|---|---|---|---|---|
| Regulation of TP53 Activity | 4 | 4,4E-03 | ENSG00000183955, ENSG00000181929, ENSG00000167491, ENSG00000012048 | 11,42 |
| Selective autophagy | 3 | 0,02 | ENSG00000188554, ENSG00000181929, ENSG00000109332 | 15,16 |
| Macroautophagy | 3 | 0,04 | ENSG00000188554, ENSG00000181929, ENSG00000109332 | 9,51 |
| Transcriptional Regulation by TP53 | 4 | 0,04 | ENSG00000183955, ENSG00000181929, ENSG00000167491, ENSG00000012048 | 5,09 |
| Autophagy | 3 | 0,04 | ENSG00000188554, ENSG00000181929, ENSG00000109332 | 8,62 |
| SUMOylation | 3 | 0,06 | ENSG00000079387, ENSG00000172262, ENSG00000012048 | 7,38 |

**ADHD & SA**

| | | | | |
|---|---|---|---|---|
| Regulation of TP53 Activity | 3 | 3,9E-03 | ENSG00000183955, ENSG00000117020, ENSG00000181929 | 26,77 |
| Alcoholic liver disease | 3 | 0,01 | ENSG00000117020, ENSG00000168036, ENSG00000181929 | 19,75 |

| | | | | |
|---|---|---|---|---|
| VEGFR2 mediated vascular permeability | 2 | 0,02 | ENSG00000117020, ENSG00000168036 | 99,09 |
| Transcriptional Regulation by TP53 | 3 | 0,02 | ENSG00000183955, ENSG00000117020, ENSG00000181929 | 11,94 |
| Salmonella infection | 3 | 0,02 | ENSG00000117020, ENSG00000168036, ENSG00000111540 | 11,33 |
| Infectious disease | 4 | 0,02 | ENSG00000117020, ENSG00000168036, ENSG00000126091, ENSG00000111540 | 5,25 |
| Generic Transcription Pathway | 4 | 0,03 | ENSG00000183955, ENSG00000117020, ENSG00000168036, ENSG00000181929 | 4,54 |
| Transcriptional and post-translational regulation of MITF-M expression and activity | 2 | 0,04 | ENSG00000117020, ENSG00000168036 | 46,35 |
| RNA Polymerase II Transcription | 4 | 0,04 | ENSG00000183955, ENSG00000117020, ENSG00000168036, ENSG00000181929 | 4,13 |
| TP53 Regulates Metabolic Genes | 2 | 0,05 | ENSG00000117020, ENSG00000181929 | 34,62 |
| RAB GEFs exchange GTP for GDP on RABs | 2 | 0,05 | ENSG00000117020, ENSG00000111540 | 31,93 |
| Endometrial cancer | 2 | 0,05 | ENSG00000117020, ENSG00000168036 | 32,14 |
| Membrane Trafficking | 3 | 0,06 | ENSG00000117020, ENSG00000181929, ENSG00000111540 | 6,65 |
| Longevity regulating pathway - multiple species | 2 | 0,06 | ENSG00000117020, ENSG00000181929 | 30,59 |
| VEGFA-VEGFR2 Pathway | 2 | 0,06 | ENSG00000117020, ENSG00000168036 | 29,62 |

| | | | | |
|---|---|---|---|---|
| Gene expression (Transcription) | 4 | 0,06 | ENSG00000183955, ENSG00000117020, ENSG00000168036, ENSG00000181929 | 3,56 |
| Adipocytokine signaling pathway | 2 | 0,06 | ENSG00000117020, ENSG00000181929 | 27,09 |
| Signaling by VEGF | 2 | 0,06 | ENSG00000117020, ENSG00000168036 | 26,61 |
| Vesicle-mediated transport | 3 | 0,07 | ENSG00000117020, ENSG00000181929, ENSG00000111540 | 5,73 |
| Rab regulation of trafficking | 2 | 0,07 | ENSG00000117020, ENSG00000111540 | 22,99 |
| Colorectal cancer | 2 | 0,08 | ENSG00000117020, ENSG00000168036 | 21,80 |
| Longevity regulating pathway | 2 | 0,08 | ENSG00000117020, ENSG00000181929 | 21,07 |
| Prostate cancer | 2 | 0,09 | ENSG00000117020, ENSG00000168036 | 19,35 |
| MITF-M-regulated melanocyte development | 2 | 0,09 | ENSG00000117020, ENSG00000168036 | 18,54 |
| Glucagon signaling pathway | 2 | 0,10 | ENSG00000117020, ENSG00000181929 | 17,72 |
| Apoptosis | 2 | 0,10 | ENSG00000117020, ENSG00000168036 | 17,31 |
| Disease | 4 | 0,10 | ENSG00000117020, ENSG00000168036, ENSG00000126091, ENSG00000111540 | 2,98 |
| Insulin resistance | 2 | 0,10 | ENSG00000117020, ENSG00000181929 | 17,40 |
| Viral Infection Pathways | 3 | 0,10 | ENSG00000117020, ENSG00000126091, ENSG00000111540 | 4,82 |

**SCZ & T2D**

| | | | | |
|---|---|---|---|---|
| Parkinson disease | 8 | 0,02 | ENSG00000168653, ENSG00000159199, ENSG00000164305, ENSG00000108671, ENSG00000186868, ENSG00000126214, ENSG00000132388, ENSG00000145335 | 3,04 |
| Notch signaling pathway | 4 | 0,02 | ENSG00000162736, ENSG00000159692, ENSG00000196498, ENSG00000078747 | 6,63 |
| Chromatin organization | 8 | 0,03 | ENSG00000183955, ENSG00000170291, ENSG00000172977, ENSG00000120071, ENSG00000196498, ENSG00000070061, ENSG00000167491, ENSG00000127663 | 2,74 |
| Chromatin modifying enzymes | 8 | 0,03 | ENSG00000183955, ENSG00000170291, ENSG00000172977, ENSG00000120071, ENSG00000196498, ENSG00000070061, ENSG00000167491, ENSG00000127663 | 2,74 |
| Alzheimer disease | 9 | 0,03 | ENSG00000168653, ENSG00000159593, ENSG00000162736, ENSG00000159199, ENSG00000164305, ENSG00000108671, ENSG00000186868, ENSG00000126214, ENSG00000145335 | 2,37 |
| Antigen processing: Ubiquitination & Proteasome degradation | 8 | 0,04 | ENSG00000164068, ENSG00000153827, ENSG00000108671, ENSG00000109332, ENSG00000159202, ENSG00000078747, ENSG00000154359, ENSG00000132388 | 2,55 |

| | | | | |
|---|---|---|---|---|
| Pathways of neurodegeneration - multiple diseases | 10 | 0,04 | ENSG00000101460, ENSG00000168653, ENSG00000159199, ENSG00000197653, ENSG00000164305, ENSG00000108671, ENSG00000186868, ENSG00000126214, ENSG00000132388, ENSG00000145335 | 2,13 |
| Class I MHC mediated antigen processing & presentation | 9 | 0,04 | ENSG00000170458, ENSG00000164068, ENSG00000153827, ENSG00000108671, ENSG00000109332, ENSG00000159202, ENSG00000078747, ENSG00000154359, ENSG00000132388 | 2,29 |
| Synthesis of active ubiquitin: roles of E1 and E2 enzymes | 3 | 0,04 | ENSG00000109332, ENSG00000159202, ENSG00000132388 | 8,83 |
| Ubiquitin mediated proteolysis | 5 | 0,05 | ENSG00000153827, ENSG00000109332, ENSG00000159202, ENSG00000078747, ENSG00000132388 | 3,62 |
| Signaling by MST1 | 2 | 0,05 | ENSG00000164078, ENSG00000173531 | 37,69 |
| Programmed Cell Death | 6 | 0,06 | ENSG00000170458, ENSG00000164305, ENSG00000108671, ENSG00000078747, ENSG00000186868, ENSG00000088808 | 2,84 |
| Amyotrophic lateral sclerosis | 8 | 0,07 | ENSG00000101460, ENSG00000108559, ENSG00000168653, ENSG00000159199, ENSG00000197653, ENSG00000164305, ENSG00000108671, ENSG00000126214 | 2,22 |

| | | | | |
|---|---|---|---|---|
| Signaling by Rho GTPases, Miro GTPases and RHOBTB3 | 13 | 0,08 | ENSG00000002822, ENSG00000242247, ENSG00000047932, ENSG00000159314, ENSG00000089639, ENSG00000172757, ENSG00000151725, ENSG00000166165, ENSG00000123329, ENSG00000126777, ENSG00000103415, ENSG00000126214, ENSG00000278259 | 1,67 |
| CDC42 GTPase cycle | 5 | 0,08 | ENSG00000089639, ENSG00000242247, ENSG00000123329, ENSG00000126777, ENSG00000159314 | 3,00 |
| Apoptosis | 5 | 0,10 | ENSG00000170458, ENSG00000164305, ENSG00000108671, ENSG00000186868, ENSG00000088808 | 2,84 |

**SCZ & CT**

| | | | | |
|---|---|---|---|---|
| Fructose and mannose metabolism | 2 | 0,05 | ENSG00000138030, ENSG00000178802 | 35,86 |
| Tryptophan metabolism | 2 | 0,06 | ENSG00000119689, ENSG00000171097 | 29,03 |
| Asparagine N-linked glycosylation | 3 | 0,09 | ENSG00000238083, ENSG00000140506, ENSG00000178802 | 5,46 |
